# Contextual DNA methylation QTLs link adversity responsivity to immune changes and evolutionary trade-offs

**DOI:** 10.64898/2025.12.04.25341605

**Authors:** Darina Czamara, Lucas Casten, Dave Juntilla, Aayah Nounu, A. Luise Weihs, Mira Erhart, Lisa Maier, Miriam Leskien, Johanna Tuhkanen, Aline D. Scherff, Jonas Hagenberg, Vera N. Karlbauer, Sandra Van der Auwera, Tanja M. Brückl, Become Study Team, Johannes Kopf-Beck, Samy Egli, Optima Study Team, Henry Völzke, Uwe Völker, Antoine Weihs, Alexander Teumer, Juliane Winkelmann, Holger Prokisch, Janine Knauer-Arloth, Lisa Feldmann, Lucas Miranda, Bertram Müller-Myhsok, Andrew H. Miller, Ferdinand Hoffmann, Sonja Entringer, Claudia Buss, Sibylle M. Winter, Louise Arseneault, Karen Sugden, Benjamin S. Williams, Stefan Sprinckmoller, Terrie E. Moffitt, Helen L. Fisher, Avshalom Caspi, Christine M. Heim, Marie Standl, Jari Lahti, Katri Räikkönen, Ellen Greimel, Gerd Schulte-Körne, Hans J. Grabe, Annette Peters, Melanie Waldenberger, Elisabeth B. Binder

## Abstract

Childhood adversity (CA) is one of the strongest risk factors for psychiatric but also numerous medical disorders, yet only a subset of exposed individuals develops these outcomes. Gene-by-environment interactions have long been proposed to account for this variability, but identifying the specific variants involved has proven difficult in studies using clinical endpoints. We therefore mapped interaction effects between genetic variants and CA at the molecular level, testing for contextual methylation quantitative trait loci (contmeQTLs) in peripheral blood from 3,471 adolescents and adults across six cohorts. Meta-analysis identified 5,120 contmeQTLs, >99% acting in *cis*, of which 20% were already detectable in three independent childhood cohorts (N=780), indicating that these effects emerge in temporal proximity to exposure. ContmeQTLs were enriched for immune-related pathways, particularly the major histocompatibility (MHC)/human leukocyte antigen (HLA)-complex, were associated with altered expression of HLA and other immune regulators, and captured disease risk beyond that explained by baseline meQTLs. A polygenic interaction responsivity score, constructed from contmeQTL variants, predicted both a larger acute transcriptional response to infection and a greater shift towards a pro-inflammatory plasma profile in CA-exposed adults, including in UK Biobank. Using ancient DNA, we found that these responsive alleles have been under positive selection, identifying molecular responsivity to adversity as an evolutionarily shaped trait whose benefit under acute challenge is accompanied by liability to immune-mediated and psychiatric disease.

## Introduction

Exposure to repeated, chronic stressful and adverse life events, especially early in life (childhood adversity (CA)), has been associated with long lasting and pervasive health consequences, with increased pleiotropic disease risk reported across psychiatric disorders and a number of other conditions, such as metabolic, cardiovascular, autoimmune and neurodegenerative disorders ^1–4^ and reduced life expectancy ^5^. Lasting biological sequelae of CA have been reported for multiple tissues, including effects on brain structure and function ^6–8^ and other organ systems ^4^. The impact of CA on the immune system has been of particular interest, as it may, at least in part, mediate increased risk for psychiatric as well as physical disorders, possibly representing a common etiological pathway underlying some of the multifaceted downstream health consequences of CA ^2^.

Data gathered from preclinical, clinical, and epidemiological studies indeed support lasting consequences of CA exposure on immune function, reflected in increased chronic inflammation and altered inflammatory responses ^4,9,10^. Chronically altered immune profiles, especially pro-inflammatory ones, in turn have been linked to several disorders for which risk has been shown to be increased by CA exposure, including major depression, cardiovascular and metabolic disease and neurodegenerative disorders ^11–14^.

However, the downstream consequences of CA on biological and health outcomes show large inter-individual variability ^15–17^ and are likely driven by a complex interplay of genetic and environmental components. Understanding the factors underlying this response variability has promise for uncovering relevant mechanisms of risk and resilience, biomarkers and novel therapeutic targets.

In order to uncover the genetic variants and relevant pathways underlying this variability, we need to consider gene×environment (G×E) interplay, where an individual’s genetic makeup influences their response to environmental exposures. However, individual loci involved in this interplay have been difficult to map, particularly when examining broad, distal phenotypes. This is likely due to the complexity and variability of the outcomes and the environmental risk factors, as well as the large sample sizes required to achieve sufficient statistical power ^18,19^.

Context-dependent quantitative trait locus (QTL) analysis for molecular outcomes has been proposed as a more tractable way to identify G**×**E interplay. Previous studies have successfully identified such contextual QTLs on the level of gene expression and chromatin accessibility, exploring cell-type, disease risk, pharmacological and immune stimuli as the variable contexts, among others ^20–26^. The studies converge on the fact that contextual QTLs are enriched for disease-associated variants.

For CA, epigenetic mechanisms could represent such an intermediate molecular phenotype for molecular QTL mapping. Epigenetic mechanisms, including DNA methylation (DNAm), have been proposed to mediate the lasting biological embedding of CA, supported by consistent evidence across human and animal studies ^27–31^. We and others have previously shown that G**×**E interplay, including for adversity, is relevant for predicting DNAm, with the variance in DNAm explained best by a combination of G and early adversity ^32–34^.

In contrast to the often negative health outcomes of chronic or misdirected immune activation ^35^, and the lasting consequences of CA exposure onto this ^4^, a functional and responsive immune system is essential for survival. Analyses from ancient DNA have illustrated this trade-off at allelic resolution ^36,37^. The clearest example is multiple sclerosis: risk allele frequencies in immune-related genes rose and underwent positive selection in Europe, probably driven by novel pathogenic challenges accompanying changes in diet, lifestyle and population density ^38^. Alleles that strengthened acute host defense persist today as risk alleles for immune-mediated disease raising the question of whether similar mechanisms could be at play for genetic variants moderating lasting epigenetic impact of CA in immune cells.

To unpack these questions, we started from established blood meQTLs ^39^ and tested for CA-dependent changes in the direction or strength of the genotype–DNAm association in peripheral blood samples of over 4,000 individuals across 9 different cohorts. We then annotated the identified contextual meQTLs functionally, with respect to disease risk as well as immune response in several additional cohorts and experimental settings, including UK Biobank. Finally, we leveraged data from more than 15,000 West Eurasian individuals spanning the past 10,000 years ^40^ asking whether molecular responsivity to CA has been under directional selection.

## Methods

Details on included cohorts, phenotype assessment and omics profiling are provided in the **Supplemental Methods**. Here, we give a brief overview on sample sizes and omics data used.

The authors used Claude Sonnet 5.5 for assistance with code development, identification of relevant literature and language editing. All code was reviewed and tested by the authors and all references were verified by the authors, who take full responsibility for the content. The graphical abstract was created using bioRender.

### Statistical analysis

A graphical overview of statistical analyses is provided in **Figure S1.** Meta-analysis across adult/adolescent cohorts and annotation of significant contextual meQTLs

Briefly, in the initial 6 cohorts (ALSPAC, BeCOME, BioMD-Y, KORA, OPTIMA and SHIP) we tested for contextual methylation quantitative trait loci (contmeQTLs) using linear regression models with the specific CpG-site as outcome and the interaction of CA and the specific SNP-genotype (coded as 0-1-2) as predictor, covarying for important confounders (age, sex, genetic PCs, estimated cell types, CpG cg05575921 indicative of smoking status ^41^). More details about this meta-analysis, the evaluation of sex-specific and celltype-specific effects, the sensitivity analysis using a fully saturated model as well as about the overlap with variance meQTLs, enrichment for chromatin states and GO terms are given in the **Supplemental Methods**.

### Expression quantitative trait methylation (eQTM) analysis in KORA and BeCOME

In KORA, we utilized *MatrixEQTL* ^42^ version 2.3 to evaluate linear associations between gene expression and DNAm (eQTM) on the 5,115 CpGs regulated by contmeQTLs, either in *cis* or *trans.* In BeCOME, we ran eQTM analysis for the two HLA-genes which presented with significant eQTMs in KORA, *HLA-DRB5* and *HLA-DRB6* (see **Supplemental Methods**).

#### Enrichment for hits associated with complex traits

To evaluate if contmeQTL SNPs and CpG-sites are associated with complex traits, we used data from the EWAS catalogue, the MRC IEU OpenGWAS platform as well as results from a Mendelian Randomization (MR) analysis performed by Richardson et al. ^43^. For each trait domain, we tested if the rate of associations for contmeQTLs SNPs/CpGs was higher as compared to the rate of associations for non-contmeQTLs SNPs/CpGs using Poisson rate tests (see **Supplemental Methods**).

#### Meta-analysis across adult/adolescent and childhood cohorts

To test which of the 5,120 significant contmeQTLs were already present during childhood, we used data from three cohorts with available genotype and DNAm data in children (GLAKU, LISA south and Kids2Health). We reran the meta-analysis across all nine cohorts. A contmeQTL was defined as replicated if (1) the meta-analysis p-value across the nine cohorts was lower as compared to the meta-analysis across the six initial cohorts and (2) the contmeQTL survived FDR-correction at 0.05.

#### Construction of polygenic interaction responsivity score (PIRS)

For the construction of a polygenic interaction responsivity score (PIRS), we used all SNPs identified as contmeQTLs. Details on the construction of the score are given in the **Supplemental Methods**, the score weights are given in Table **S13**. The score is based on the sum of responsive alleles and represents the molecular responsiveness, measured as change in DNAm, to the interaction of genetic variants and CA.

#### Association of PIRS with inflammatory markers in UK Biobank and E-Risk

In UK Biobank, we tested the association of the PIRS with the inflammatory marker profile of 31 markers using multivariate linear regression with PIRS and CA status as predictors, followed by a permutation-based paired Wilcoxon-test to compare the standardized effects of the PIRS on each pro-inflammatory marker separated by exposure groups (see **Supplemental Methods** for further details). In E-Risk DZ twins, we similarly assessed the association of the PIRS in context of CA with CRP, IL-6 and suPAR using linear mixed models with the family-ID as random effect.

#### Association of PIRS with viral infection in PBMCs

To test whether contmeQTLs predicted individual differences in acute immune response, we applied the PIRS to an experiment examining how genetics and transcriptomics relate to immune response by Randolph et al. ^44^. Briefly, this study generated imputed genotypes for 89 samples with paired since-cell RNA-sequencing on blood samples exposed to a mock condition or the influenza A virus. We computed an IFN response score and associated it with the PIRS (see **Supplemental Methods**).

#### Evolutionary analyses on PIRS

To understand the evolutionary history of contmeQTLs, we looked for evidence of recent positive selection using a large dataset of genome-wide genotypes and selection estimates from a study on > 15,000 humans from the past ∼10,000 years ^40^. We tested if alleles included in the PIRS have been significantly positively selected for, if a higher PIRS was associated with longer survival, whether positively selected contmeQTLs were associated with increased genetic risk signal for autoimmune or neuropsychiatric traits and characterized the biological pathways influenced by positive selection at contmeQTLs (see **Supplemental Methods**).

## Results

We analyzed potential contextual methylation quantitative trait loci (contmeQTLs) in peripheral blood with regard to childhood adversity (CA) in six independent cohorts of adolescents and adults with an overall sample size of 3,471 individuals including 21% with exposure to significant CA (see **Table 2**). An overview of the research question and main results is depicted in the **graphical abstract**.

**Table 1:**
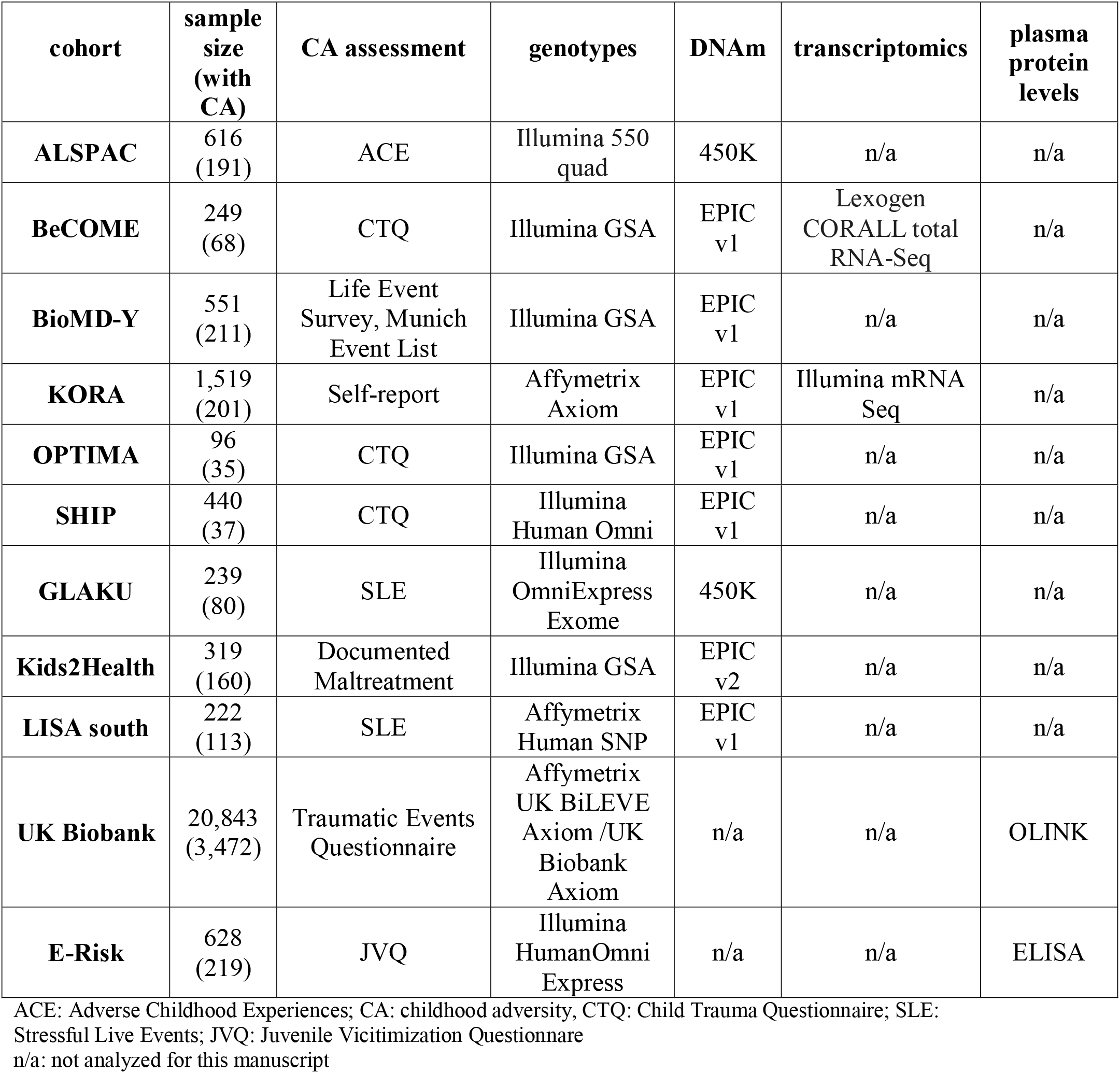
Overview of cohorts, early life adversity assessment and molecular phenotypes.

**Table 2:**
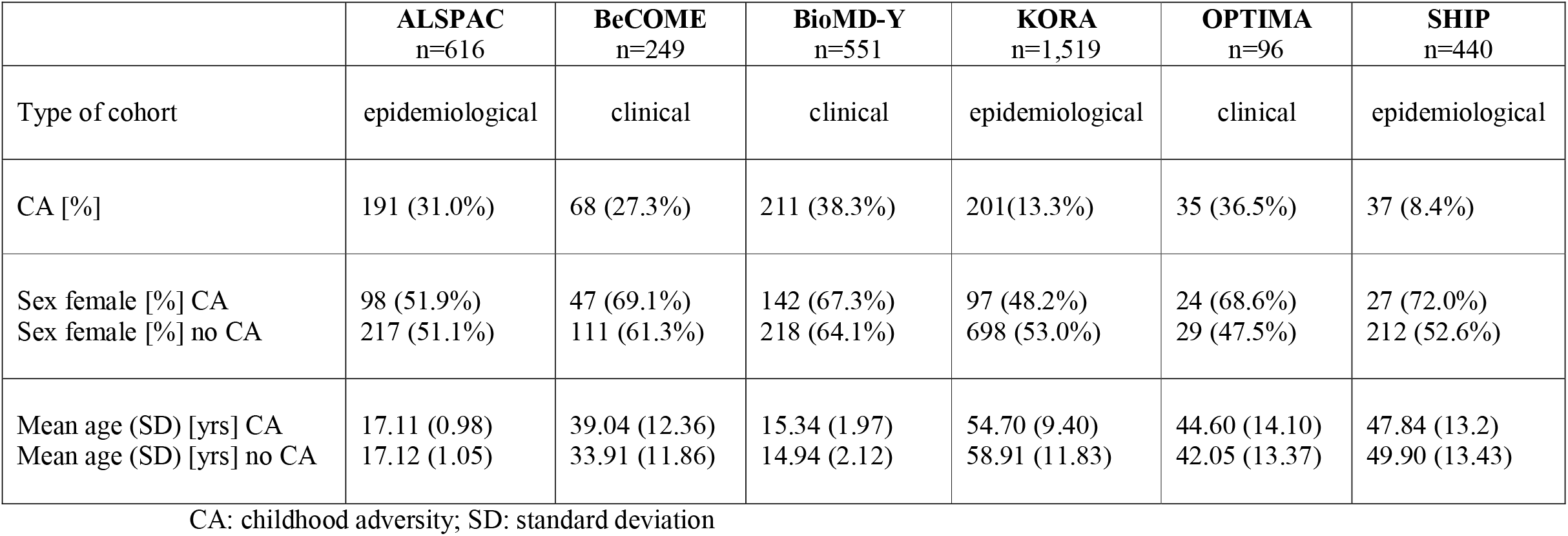
Demographics of initial cohorts.

### Identification of robust sex-consistent contmeQTLs in adolescent and adult cohorts

We restricted our search to already established meQTLs in peripheral blood. The GoDMC consortium analyzed blood meQTLs in over 30,000 individuals from European ancestry ^39^ and identified 66,577,913 significant *cis* meQTLs (consisting of 6,316,998 unique SNPs and 183,207 unique CpG-sites, p< 1×10^-08^) and 3,657,300 significant *trans* meQTLs (consisting of 893,991 unique SNPs and 19,147 unique CpGs, p<1×10^-14^), based on a *cis*-window of 1 MB. We used this list of SNP-CpG combinations and first tested for contmeQTLs with CA in each cohort (ALSPAC, BeCOME, BioMD-Y, KORA, SHIP and OPTIMA) separately (see **Methods**). We then combined results across cohorts using random-effects meta-analysis with 743 with CA and 2,728 individuals without CA in total. Overall, 5,090 contmeQTLs in *cis* and 30 contmeQTLs in *trans* survived multiple testing correction (see **Figure 1A** and **B** and **Tables S1** and **S2**). ContmeQTLs were sex-consistent, without strong cell-type specific effects and in general the SNP and the respective CpG-site were further apart from each other in these pairs as compared to all tested non-significant combinations (see **Supplementary Results 1**).

**Figure 1.**
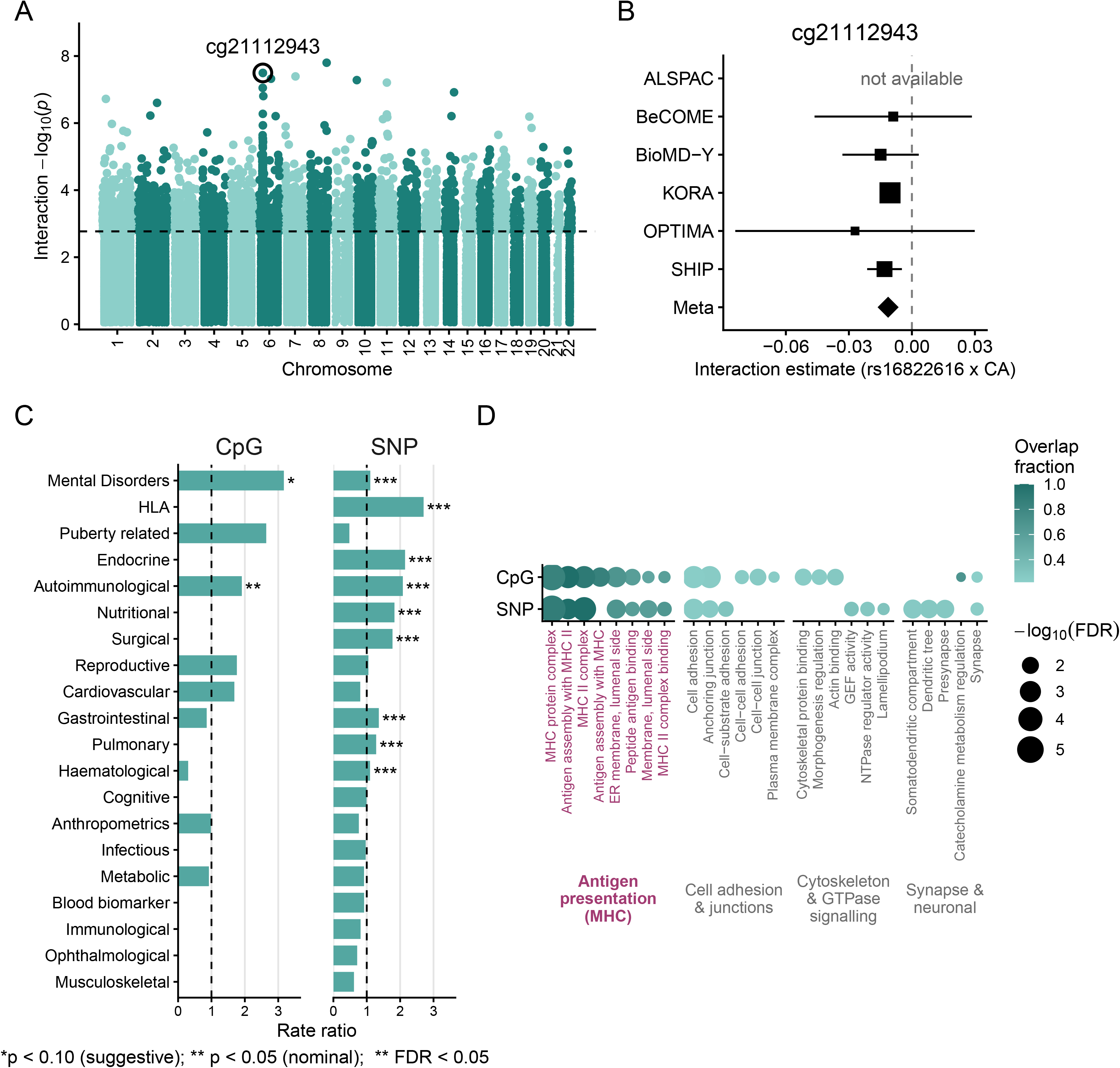
**(A)** Manhattan plot of contextual meQTLs: chromosomal position (based on the CpG-site) is depicted on the x-axis, meta-analysis interaction p-values for SNP**×**CA on the y-axis, based on the meta-analysis across 6 cohorts (N=3,471) and adjusted for genetic sex, age, cell type proportion, genetic PCs and smoking. For each CpG-site the p-value for the best combination is plotted. The dashed line marks the p-value threshold corresponding to FDR of 0.05. The CpG-site within the MHC region with the lowest interaction p-value for SNP**×**CA is highlighted. **(B)** Forest plot of the interaction of SNP**×**CA on DNAm for top hit in the MHC region: rs16822616 interacting with CA on cg21112943. For each cohort, the interaction estimates, adjusted for genetic sex, age, cell type proportion, genetic PCs and smoking, represented by a square inversely proportional in size to the standard error as well as the 95% confidence interval are depicted. The dashed line represents an interaction effect of 0. The meta-analysis interaction effect is depicted by a diamond. The specific combination of rs16822616 and cg21112943 was available in five out of the six cohorts (BeCOME, BioMD-Y, KORA, OPTIMA and SHIP) and interaction effect directions were consistent across the cohorts. **(C)** Rate ratios comparing the rate of GWAS hits and MR hits among contextual meQTLs with the rate among noncontextual meQTLs (based on exact Poisson test) across different domains. The dashed line at 1 indicates no enrichment. Enrichment was based on MR-results by Richardson et al. (2018) as well as on GWAS hits from the IEU OpenGWAS catalogue. **(D)** Enrichment of GO terms among genes mapped to CpG-sites and SNPs involved in contmeQTLs. The color gradient represents the overlap of the genes mapped by closest distance to SNPs/CpGs relative to all genes present in the specific GO term, the size of the dot the FDR-corrected p-value. Only results significant after multiple testing correction in each analysis (CpGs and SNPs) are shown. GO terms implicated in the MHC complex/antigen binding are highlighted.

In addition, we performed further sensitivity analyses including BMI as covariate as well as running the fully saturated model, i.e., including the interaction term of every covariate with the respective SNP and CA. Interaction effects estimates were highly concordant with those from the primary analyses (mean correlation across cohorts r=0.97, see **Supplementary Methods** and **Supplementary Results 2**).

#### ContmeQTLs are enriched for variance meQTLs and enhancers

Since gene-environment interactions can increase phenotypic variability in general and variability of DNAm in particular ^45^, we hypothesized that contmeQTLs might be enriched for variance meQTLs (vmeQTLs), i.e. SNPs associated with the variability of DNAm. Indeed, both SNPs acting as contmeQTLs (‘contmeQTL SNPs’) and CpGs regulated by contmeQTLs (‘contmeQTL CpGs’) were enriched for vmeQTLs as reported in Ek et al. ^46^ (p=0.006, based on 1,000 permutations for SNPs and p=0.001, 1,000 permutations for CpG sites, respectively).

To evaluate the potential gene regulatory function of contmeQTL SNPs and contmeQTL CpGs, we performed enrichment analyses for chromHMM marks. We found significant enrichment for enhancers, among other regulatory sites (see **Supplementary Results 3**).

#### ContmeQTLs overlap with GWAS hits

We next checked if contmeQTL SNPs and CpGs are associated with complex traits. The proportion of overlap with GWAS hits was significantly larger for contmeQTL SNPs (overlap of 44.7%, n=3,778 SNPs out of 8,425 unique contmeQTL SNPs) as compared to all tested meQTL SNPs (overlap of 36.8%, n=61,740 SNPs out of 167,600 unique tested SNPs; p=3.54×10^-23^, rate ratio=1.23, Poisson rate test). We not only found a higher degree of overlap, but also significant differences with regards to domains the GWAS hits mapped to: contmeQTL SNPs mapped significantly more often to the domains ‘Autoimmune immunological disease’, ‘Endocrine’, ‘Gastrointestinal’, ‘Haematological’, ‘HLA’, ‘Mental Disorders’, ‘Nutritional’, ‘Pulmonary’ and ‘Surgical’ compared to all tested meQTL SNPs (see **Figure 1C**, **Supplementary Results 4**, **Tables S3** and **S4**). The relative proportion of CpG-sites overlapping with EWAS-hits from the EWAS catalogue did not significantly differ between all tested meQTLs CpGs and contmeQTL CpGs (details on the EWAS overlap are given in **Supplementary Results 4** and **Tables S5** and **S6**).

#### CpG-sites regulated by contmeQTLs are potentially causally related to mental health

We next employed Mendelian Randomization (MR) to check for potential causal relations with disease traits. Therefore, we leveraged the systematic MR-analysis by Richardson et al. ^43^ that assessed the relationship between CpG-sites and 139 complex traits using blood meQTL SNPs as instruments. We observed a significantly higher overlap: overall, 0.33% of all tested CpG-sites overlapped with CpG-sites identified as causal on any complex traits in ^43^, whereas 0.6% of contmeQTL CpGs overlapped with this list (p=3.39×10^−0^^4^, rate ratio=1.88, Poisson rate test). We also found differences with regard to domains the complex traits mapped to (see **Tables S7** and **S8**), with significantly stronger enrichment for ‘Autoimmune immunological disease’ (e.g., “Rheumatoid arthritis”, rate ratio =1.91, p=4.90×10^-02^) and a tendency towards stronger enrichment for Mental Disorders’ (e.g., “Schizophrenia”, rate ratio= 3.17, p=5.10×10^-02^). Additionally, we found higher overlap for the trait domains ‘Cardiovascular’ (e.g., “Coronary artery disease”), ‘Puberty related’ (i.e. “Age at menarche”) and ‘Reproductive’ (i.e. “Age at menopause”, see **Figure 1C**) over all blood meQTLs, reflecting in part the GWAS enrichments reported above.

#### ContmeQTLs are enriched in pathways implicated in the major histocompatibility (MHC)-protein complex and associated with gene expression of MHC genes

Enrichment for GO terms highlighted several pathways implicated in the major histocompatibility complex (MHC)/human leukocyte antigen (HLA)-complex (located on chr6:28,477,797-33,448,354, based on hg19, see **Figure 1D**), reflecting the GWAS overlap with HLA (see **Figure 1D**). Genes mapping to contmeQTL CpGs and contmeQTL SNPs were significantly enriched for the following GO terms: MHC-protein-complex, peptide antigen assembly with MHC-class-II-protein complex, MHC-class-II-protein-complex, peptide antigen assembly with MHC-protein complex, luminal side of endoplasmic reticulum (ER) membrane, peptide antigen binding, lumenal side of membrane, and MHC-class-II-protein-complex binding. More details on these enrichments are given in **Supplementary Results 5**. The link to the MHC could further be strengthened by expression quantitative trait methylation (eQTM) analyses in KORA and BeCOME (N = 1519 and 179, respectively) with significant associations between contmeQTL CpG-sites and gene expression of MHC genes as well as other genes relevant to immune regulation (see **Supplementary Results 6** and **Table S9)**.

We next investigated if any markers showed associations across all of these layers underscoring the complex mechanisms which might underlie regulation by contmeQTLs and identified 16 contmeQTL CpGs that overlapped with eQTMs, had been associated with complex traits in the MR-analysis ^43^ and overlapped with EWAS hits as well as GWAS hits for complex traits (see **Supplementary Results 7, Figure S2** and **Table S10**).

#### ContmeQTLs already act in childhood cohorts

Next, we wanted to establish if contmeQTLs identified in adolescents and adults already show significant moderation by CA early in life. For this, we used data from three independent childhood cohorts: GLAKU, Kids2Health and LISA including 780 children aged 3-13 years (see **Table 3**). Information on CA was available for all cohorts using different instruments (see **Methods**). Of the initial 5,120 (5,090 in *cis*, 30 in *trans*) contmeQTLs, n=1,061 (i.e., 20%, with 1,052 in *cis* and 9 in *trans*) could be replicated in the childhood cohorts (see **Tables S11** and **S12**). This overlap is not only significantly more than expected by chance (p<2.2×10^-16^, Binomial test, assuming a of 5% significant results by chance) but also consistent in effect direction with nearly all contmeQTLs from the initial meta-analysis having the same interaction effect direction also in the meta-analysis across the childhood cohorts (only 4 contmeQTLs differ in their effect direction).

**Table 3:** Demographics of replication cohorts.

|  | **GLAKU**<br>n=239 | **Kids2Health**<br>n=319 | **LISA**<br>n=222 |
|---|---|---|---|
| Type of cohort | epidemiological | selected | epidemiological |
| CA [%] | 80 (33.5%) | 160 (50.1%) | 113(50.9%) |
| Sex female [%] CA<br>Sex female [%] no CA | 39 (48.8%)<br>84 (52.8%) | 73 (45.6%)<br>68 (42.8%) | 49 (43.4%)<br>45 (41.3%) |
| Mean age (SD) [yrs] CA<br>Mean age (SD) [yrs] no CA | 12.39 (0.47)<br>12.33 (0.52) | 8.31 (2.19)<br>7.94 (2.00) | 6.07 (0.18)<br>6.07 (0.11) |

CA: childhood adversity; SD: standard deviation

Compared to the enrichments observed in adolescents and adults, these sites were even more enriched for MHC and HLA-related pathways (p-values for enrichment over adult/adolescent sites from 1.63×10^−06^ to 2.71×10^−05^, see **Supplementary Results 8**). This suggests, that contmeQTLs, including within MHC/HLA loci, are already apparent in early life, proximal to the exposure.

#### A polygenic interaction responsivity score is associated with higher inflammation and immune response

Next, we computed a polygenic interaction responsivity score (PIRS), based on the 3,475 LD-clumped SNPs acting as contmeQTLs. This score is the sum of copies of responsive alleles, i.e., alleles associated with higher absolute DNAm changes in the context of CA exposure (see **Methods** and **Table S13**). In UK Biobank, the PIRS was significantly associated with a pro-inflammatory shift based on differences in the plasma levels of 31 pro-inflammatory markers from the OLINK panel (p=0.004 joint Wald-test using a robust covariance matrix). While we found no significant interaction effect of CA and the PIRS on the multi-marker levels, effect sizes of the PIRS on the individual pro-inflammatory markers were significantly higher in the CA group (p=0.048, permutation-based Wilcoxon-test, see **Methods** and **Figure 2A**).

**Figure 2.**
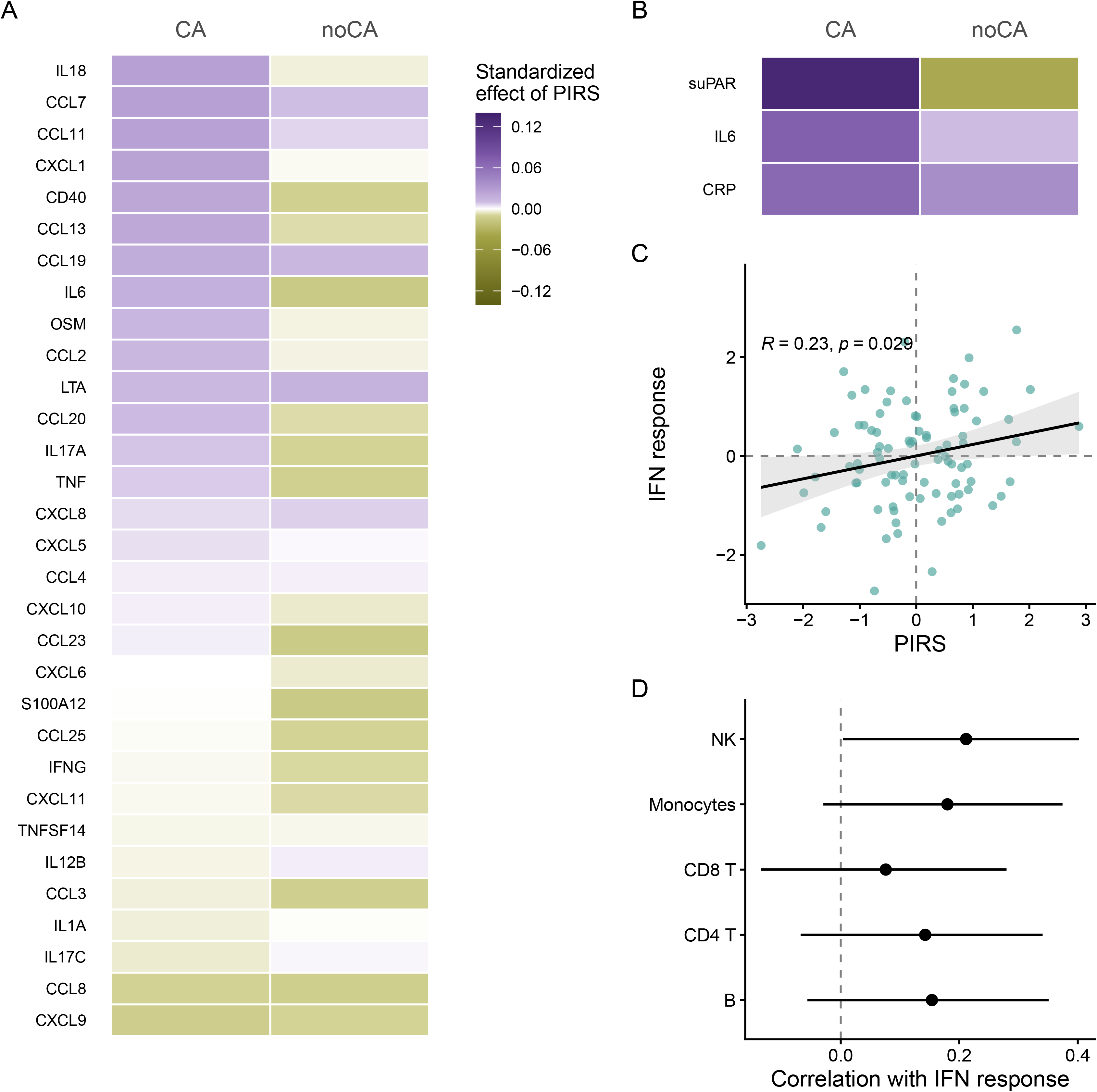
**(A)** For each tested plasma protein marker in the UK Biobank, the standardized effect of PIRS on the specific plasma protein marker, adjusted for genetic sex, age, BMI and genetic PCs, is depicted as color gradient. The sample was stratified by exposure group, the standardized effect size for CA exposed (n=3,472) is depicted in purple, for non-CA exposed (n=17,371) in olive. The p-value for the joint Wald-test of the PIRS across all 31 pro-inflammatory plasma protein markers, adjusted for genetic sex, genetic PCs, age, BMI and CA exposure and using a robust covariance matrix was 0.004. The p-value for a Wilcoxon-test for significant differences between PIRS effect sizes in the CA and no CA groups, based on 10,000 permutations, was 0.048. **(B)** The standardized effect of PIRS on available plasma protein markers in DZ twins age 18 years from the E-Risk cohort, adjusted for genetic sex, BMI and genetic PCs, is depicted, based on the same color gradient as in (A). The sample was stratified by exposure group, the standardized effect size for CA exposed (n=219) is depicted in purple, for non-CA exposed (n=409) in olive. In the exposed group, a higher PIRS was significantly associated with higher suPAR levels (p=0.014, p_FDR_=0.042). **(C)** Scatterplot of PIRS and IFN transcriptional response after influenza infection in pseudo-bulk PBMC cells from 89 males from Randolph et al. (2021). The regression line is based on a linear model of PIRS onto IFN transcriptional response, adjusting for the same covariates as used in Randolph et al., i.e., cell type proportions, PCs, ethnicity, age, age^2^ and baseline IFN levels. The shaded area represents the 95% CI of the linear fit. **(D)** Spearman correlation (and 95% CI) of PIRS and IFN transcriptional response after influenza infection in single cell types in Randolph et al. The vertical dashed line represents no correlation. Across all cell-types, the direction of association is consistent.

Given the median age of 58 years in the UK biobank sample, participants have likely accumulated an inflammatory burden throughout their lives in addition to the exposure to CA. This is particularly relevant as inflammation tends to increase with age. We therefore investigated whether we could replicate our findings in a younger sample, where the cumulative influence of these factors would be less pronounced and CA exposure itself occurred closer to the assessment of the inflammatory markers. For this, we used data from dizygotic twins aged 18 years from the E-Risk study, 35% having experienced maltreatment and other adversities in their childhood or adolescence. Three inflammatory plasma markers (CRP, IL6 and suPAR) were available. Similar to our findings in UK Biobank, PIRS regression effects in exposed individuals were higher as compared to non-exposed individuals for all three markers (see Figure **2B**). Additionally, a higher PIRS was significantly associated with higher suPAR levels in exposed individuals (p_CA_=0.014, p_FDR_=0.042, one-sided test) but not in non-exposed individuals (p_noCA_=0.23). Furthermore, we observed a significant interaction between exposure status and the PIRS on suPAR levels in the combined sample (p=0.03).

Given the observed association between PIRS and pro-inflammatory markers, we next tested whether the PIRS predicted individual differences in an acute experimental immune response. We utilized paired single cell RNA-seq data from peripheral blood mononuclear cells (PBMCs) exposed to a mock condition or influenza A infection for 6 hours in n=89 genotyped individuals (235,161 cells) based on publicly available data from Randolph ^44^. We computed a transcriptional response score, measuring the strength of the immune response to infection, namely increased interferon pathway gene expression, as described in ^44^. The PIRS was significantly positively associated with the magnitude of the immune response (p=0.029, see **Figure 2C**) and mainly driven by NK cells (see **Figure 2D**).

#### The polygenic interaction responsivity score is associated with survival advantage with trade-offs for autoimmune and psychiatric disorders

These results suggest that contmeQTLs shape individual differences in the strength of the immediate transcriptional responses to infection as well as lasting proinflammatory changes following CA, highlighting that these CA responsive alleles might associate with survival advantages and thus possibly positive selection over human history.

To test this, we conducted a series of exploratory evolutionary analyses using data from a recent genome-wide selection scan and matched genotypes from >15,000 ancient West Eurasian individuals ^40^. The single strongest genome-wide signal was at the contmeQTL SNP rs4785704 (selection p=1.80×10LJ¹¹), for which the allele frequency of the more responsive allele was near zero until approximately 10,000 years ago and then rose steadily to ∼7% in present-day populations (see **Figure 3A**). More broadly, contmeQTLs included in the PIRS were 2.47 times more likely to be significantly positively selected than non-contmeQTLs, independent of allele frequency (Firth logistic regression OR 95% CI: 1.42-4.10, p = 0.002). This was supported by permutation analyses (see **Figure 3B** and **Supplementary Result 9**). For downstream analyses, we focused on the sum of the alleles of the 18 positively selected variants at genome-wide significance, labeled positive selection PIRS or PIRS+.

**Figure 3.**
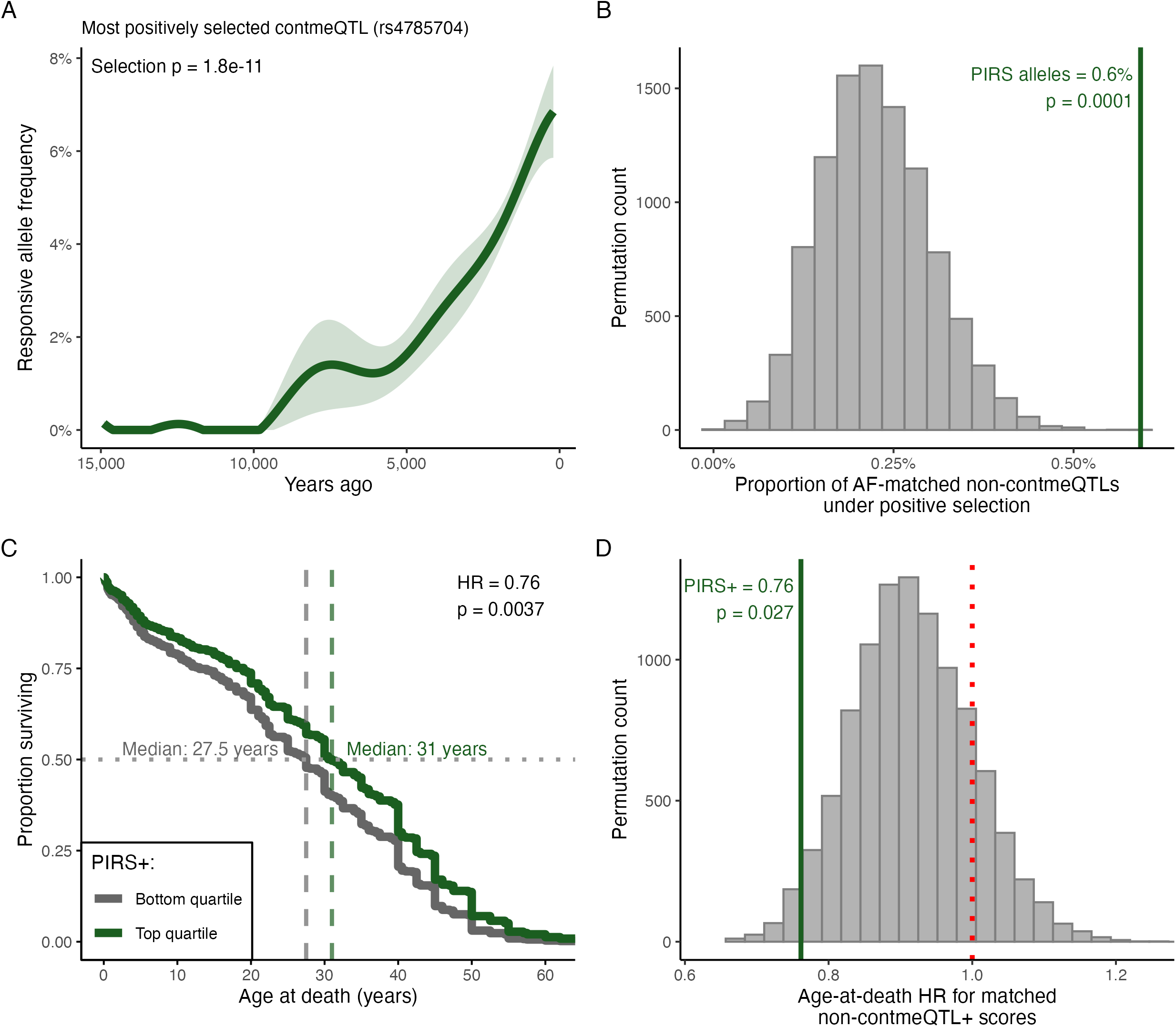
**(A)** Frequency over time of the responsive allele at rs4785704, the contmeQTL under the strongest positive selection in ancient DNA (Akbari et al., 2026). Shading indicates 95% CI for allele frequency. **(B)** The proportion of LD-pruned PIRS variants under positive selection (FDR < 0.05), indicated by the vertical dark green line, as compared to the proportion of positively selected variants in 10,000 sets of LD-pruned non-contmeQTLs, matched for allele frequency and set size to the PIRS variants. All permutations yielded a lower selection rate than the observed one, resulting in an empirical p-value of 0.0001 for positive selection of PIRS variants. **(C)** Kaplan-Meier curves of estimated age-at-death (available in 983 ancient individuals) within the top (dark green) and bottom (dark grey) PIRS+ quartiles. The Hazard Ratio (HR) from Cox regression adjusted for sample date, sex and array coverage is significantly lower in individuals presenting within the top quartile (HR=0.76, p=0.0037, median age of 31 years) as compared to individuals within the bottom quartile (median age of 27.5 years). **(D)** Age-at-death HRs for 10,000 top vs. bottom quartile scores, based on positively selected non-contmeQTLs, matched in allele frequency and set size to the original PIRS+ score. The dark green vertical line indicates the HR of the original PIRS + variants. 270 permutation-based scores yielded a lower HR than the original PIRS+ variants, resulting in an empirical p-value of 0.027 for a significantly lower HR in individuals within the top PIRS+ quartile as compared to individuals within the bottom PIRS+ quartile. The red dashed line indicates a HR of 1, i.e., no difference between the two groups.

To test whether this selection signal reflected survival advantage, we examined estimated age-at-death among N = 983 ancient individuals with imputed genotype data from the Allen Ancient DNA Resource ^40,47^ using Cox proportional-hazards regression. Individuals in the top quartile of the PIRS+ lived to a significantly older estimated age than those in the bottom quartile (median 31 vs. 27.5 years estimated age-at-death; Cox HR = 0.76, p = 0.0037, see **Figure 3C**). Using permutations analyses, we could show that the survival advantage of PIRS+ was significantly more pronounced as compared to matched positively selected but non-contmeQTLs, (see **Figure 3D** and **Supplementary Results 10**) underscoring that positively-selected contmeQTLs confer a survival advantage in ancient humans beyond what would be expected for similarly-selected variants.

Positively-selected contmeQTLs were not only broadly enriched for immune-related pathways but also carried elevated GWAS risk, most strongly for autoimmune disease (see **Figures S3A** and **B** and **Supplementary Results 11**).

Sensitivity analyses of varying the threshold for positive selection and hence the number of included SNPs, ranging from 14 to 340, indicated robustness of these findings on enrichment for positive selection, survival advantage as well as enrichment for immune-related pathways and GWAS hits (see **Supplemental Methods, Supplemental Results 12** and **Figure S4A-C**).

## Discussion

In order to map lasting epigenetic effects of CA in immune cells, we investigated meQTLs in the context of CA. We identified over 5,000 significant contmeQTLs in adolescents and adults in peripheral blood, that is meQTLs that were moderated by the exposure of CA, and around 1,000 of these were already significant in children. SNPs and CpG sites within these contmeQTLs mapped to genes that were enriched for the MHC region and eQTM analyses confirmed that differential DNAm at contmeQTL CpGs was associated with altered gene expression, including for *HLA-*genes, supporting their functional relevance. Significant enrichment of contmeQTL SNPs for GWAS hits for disease traits above meQTLs lends support for the importance of G**×**E in complex traits. This was in line with the significantly higher enrichment of contmeQTL CpGs in MR associations of blood meQTLs. In agreement with the pleiotropic risk conveyed by CA, enriched traits included mental disorders but also autoimmune disorders and asthma. This genetic responsivity, when summarized into a polygenic score, predicted both larger acute transcriptional response to a viral challenge as well as a greater shift in lasting pro-inflammatory profiles in CA-exposed participants. Interestingly, the strongest effect was observed for suPAR, a stable plasma protein released by proteolytic cleavage of membrane-bound uPAR (urokinase plasminogen activator receptor) from monocytes, neutrophils and activated lymphocytes, its circulating levels likely reflect the activation state of the immune cells in which the contmeQTLs were mapped ^48^. suPAR is also a transdiagnostic marker: higher levels predict cardiovascular disease, diabetes and mortality independently of CRP and other established risk factors ^49^, which parallels the transdiagnostic risk profile of the contmeQTL.

The fact that this molecular responsivity seemed to have beneficial health consequences in the context of acute immune challenges, such as viral infections, as well as negative health consequences in a more chronic exposure context is in line with the observation, that the responsive alleles underwent positive selection over the last 10,000 years of human history.

The results of our study support the importance of G**×**E for complex traits for which CA is an environmental risk factor. ContmeQTLs explained disease risk beyond meQTLs. While the role of G**×**E in CA-related disorders has long been proposed, as for example by the stress-diathesis model ^50^, mapping specific genes in this context has been more challenging ^51^. Genome-wide approaches have tried to explore this, with some supportive results for major depression as outcome ^52^. Overall, however, genome-wide G**×**E for CA are challenging to conduct and interpret due to the need of very large sample sizes and variability in how both CA and the diverse outcomes are reported and measured ^53–55^.

Focusing on molecular traits as proximal measures that are more reliably quantifiable may enable more robust detection of specific variants within G**×**CA. We focused on methylation QTLs, given the potential impact of CA on the epigenetic landscape and, to increase robustness, started from meQTLs discovered in the largest peripheral blood meQTLs analysis to date with over 30,000 individuals included ^39^. Using this contextual QTL approach, our analysis identified over 5,000 robust contmeQTLs with CA using random effects meta-analysis over 6 independent cohorts with over 3,400 individuals. While prior studies on contextual QTL approaches have focused on disease states, physiological differences or pharmacological challenges ^20–26^, our results show that contextual QTLs can also capture G**×**E with external, complex environments such as CA.

In the adult and adolescent cohorts, we used the Childhood Trauma Questionnaire (available in the BeCOME, OPTIMA and SHIP cohorts) ^56,57^ as well as other validated questionnaires for CA that all focus on retrospective report of abuse, maltreatment and neglect occurring before the age of eighteen. About 20% of the contmeQTLs were already significant in our three childhood cohorts (N = 780) that ranged in age from 3 to 13 years. These used caregiver/parental reports of maltreatment (Kids2Health) or parental/maternal reports of adverse life events such as divorce, severe disease, death or unemployment (LISA and GLAKU). This suggests that contmeQTLs emerge already in temporal proximity to CA exposure, and are not solely reliant on subsequent health changes. However, future studies will need to examine differential effects of developmental timing on contmeQTLs in more detail ^58,59^.

As mentioned above, CA is a complex exposure. Interestingly, we observed similar effects across epidemiological cohorts (ALSPAC, GLAKU, KORA, LISA and SHIP) as well as clinical cohorts (BeCOME, BioMD-Y and OPTIMA) and exposure assessments (retrospective self-reports of adults as well as more proximal reporting by parents or caregivers). The fact that we detected similar effects across different instruments, populations and rates of CA supports that the identified interaction effects are not specific to one type of exposure. However, additional work is needed to explore potential exposure-specific interactions in more detail. It is important to note that reports of CA may not solely reflect discrete traumatic events but likely also capture broader disadvantageous childhood environments. Adverse childhood experiences are strongly socially patterned and tend to co-occur with socioeconomic disadvantage and other adverse conditions. Consequently, one specific measure of CA such as the CTQ may partly serve as a proxy for a wider constellation of early-life adversities, including prenatal factors ^60^.

ContmeQTLs in immune cells may provide a link between CA and the reported pleiotropic long term health risk that could be mediated by the joint impact of genetic risk variants and CA on immune function. Reflecting this, CA-contmeQTLs were linked to pleiotropic disease risk both at the SNP and the CpG level, including through MR analysis from blood meQTLs. In fact, overlap with GWAS and from MR analyses indicates enrichment for mental disorders but also autoimmune/immunological disorders, including asthma and other disorders (see **Figure 1C).**

The identified contmeQTLs support lasting effects of CA and genetic susceptibility on immune regulation via epigenetic changes. This is underscored by an enrichment in genes from the MHC-complex and the fact that eQTM results show that these changes in DNAm also relate to altered expression in *HLA*-genes and other immune regulators. By computing a polygenic interaction responsivity score (PIRS) with the 3,475 LD-clumped contmeQTL SNPs, we could interrogate the combined impact of these changes on the immune system. Indeed, the PIRS, with higher values indicative of higher variance in DNAm in the context of CA, was significantly associated with a shift towards a pro-inflammatory profile in CA-exposed participants in the UK Biobank and E-Risk cohorts. This suggests that this genotype-dependent epigenetic embedding of CA is associated with a pro-inflammatory immune profile from early adulthood on, supporting the need for early detection of these states and intervention. The proposed PIRS and accompanying DNAm measures in peripheral blood could be developed into a biomarker for such CA-dependent risk.

The pleiotropic consequences of molecular responsivity reflect a more general property of the system in which we measured it. Immune activation is Janus-faced: the same response that limits infection also damages host tissue, and the genetic factors impacting immune response have been shaped by selection under conditions that differ from those of contemporary life ^35,36,38,61^. Ancient DNA studies have begun to document the consequences of this. Genetic adaptation to pathogens in post-Neolithic Europe was enriched in genes relating to host–pathogen interactions and is accompanied by an increase in the risk of inflammatory disorders ^36^. For multiple sclerosis, the risk variants in immune genes could be traced to positive selection among Pontic steppe pastoralists and subsequently in Europe, probably driven by pathogenic challenges accompanying changes in diet, lifestyle and population density ^38^. While these studies show that disease-associated immune alleles have been under selection, they can only presume the trait on which selection acted, as the phenotypes were relevant for Bronze Age pastoralists and not modern humans. ContmeQTLs provide an opportunity to address this. Responsive alleles are defined by the magnitude of the molecular response in the context of CA rather than by its long-term negative health outcomes. They thus index a molecular regulatory capacity that impacts immune function. The fact that the alleles contributing to the PIRS show evidence of positive selection, while being enriched for autoimmune and psychiatric risk in current GWAS, illustrates the trade-off of optimizing response to infection vs. damaging host tissue within a set of functional genetic variants. What appears to have been favored is the capacity for a molecular response to the environment, where environment seems to encompass viral infections as well as adverse life events. The pro-inflammatory profile and pleiotropic disease risk we observe today represents this capacity in a context in which the predominant exposures are chronic and psychosocial rather than acute and infectious.

## Conclusion

This study supports the relevance of G×E in the epigenetic embedding of CA in immune cells and proposes changes in immune regulation as one common pathway linking CA to pleiotropic disease risk. The contmeQTL CpGs were associated with altered expression of genes regulating inflammation, and the polygenic interaction responsivity score derived from these loci predicted both a larger acute transcriptional response to a viral challenge and a greater shift towards a pro-inflammatory profile in CA-exposed participants. The fact that the underlying alleles show evidence of positive selection suggests that responsivity is not primarily a deficit, but a graded trait with context-dependent consequences. What appears to have been advantageous when exposures were acute, may carry a cost where they are chronic or psychosocial, and this could contribute to the link between CA and transdiagnostic disease risk in later life. The PIRS, together with the accompanying DNAm measures in peripheral blood, could be developed into a biomarker identifying individuals who shift towards an inflammatory phenotype after exposure to CA, and who might benefit from adapted prevention and treatment approaches, as suggested for patients with CA and increased inflammatory tone ^1,62^.

## Acknowledgements

We acknowledge the valuable contributions of Christian Gieger, who sadly passed away during the course of this work. We are grateful for his input and contributions to this study. We are extremely grateful to all the families who took part in this study, the midwives for their help in recruiting them, and the whole ALSPAC team, which includes interviewers, computer and laboratory technicians, clerical workers, research scientists, volunteers, managers, receptionists and nurses.

The authors would like to thank the Psychiatric Study Center of the Max Planck Institute of Psychiatry - Dr. Norma Grandi, Anna Gossmann, Elisabeth Kappelmann, Karin Hofer, Alexandra Kocsis, Vlada Kolysnik and Gertrud Ernst-Jansen - for organizational support, biomaterial and data collection and the Biomaterial Processing and Repository Unit of the Max Planck Institute of Psychiatry - Tamara Namendorf, Božidar Novak, Marketa Reimann, Angelika Sangl - for the processing and storage of the study’s biosamples. The authors further thank Julia-Carolin Albrecht, Anastasia Bauer, Anja Betz, Alina Feichtinger, Rebekka Hirsch, Miriam El-Mahdi, Eila Mertens, Carolin Haas, Katharina Kahn, Lisa Kammholz, Sophia Koch, Christof Leininger, Anna Lorenz, Rebecca Meissner, Jessie Osterhaus, Liisbeth Pirn, Christina Rieger, Linda Schuster and Refican Yasar for their help with data collection, study management, the recruitment, and screening of BeCOME participants. The authors’ special thanks go to all study participants for participation in the BeCOME study.

We thank all the GLAKU children and their parents for their enthusiastic participation. We also thank all the research nurses, research assistants, and laboratory personnel involved in the GLAKU study.

We thank all participants for their long-term commitment to the KORA study, the staff for data collection and research data management and the members of the KORA Study Group (https://www.helmholtz-munich.de/en/epi/cohort/kora) who are responsible for the design and conduct of the study.

The authors thank all families for participation in the LISA study and the study teams for their excellent work.

The SHIP authors are grateful to Holger Prokisch and Thomas Meitinger (Helmholtz Zentrum München) for the genotyping of the SHIP-TREND-0 cohort.

We are grateful to the E-Risk study mothers and fathers, the twins, and the twins’ teachers and neighbours for their participation. Our thanks to all members of the E-Risk team for their dedication, hard work and insights.

## Data availability

Due to ethical issues and consent, datasets analyzed during the current study are not publicly available. Interested researchers can obtain a deidentified dataset after approval from the respective study boards.

## Code availability

The code for the main analyses is available at: https://github.com/darinacz/contextual_meQTLs

## Authors’ contributions

DC: study design, data analysis, data interpretation, writing of manuscript

LC: data analysis, data interpretation, writing of manuscript

DJ: data analysis, data interpretation

AN: data analysis, data interpretation

ALW: data analysis, data interpretation

ME: data analysis, data interpretation

LMa: data analysis, data interpretation

ML: data analysis, data interpretation

JT: data analysis, data interpretation

ADS: data collection, data interpretation

JH: data analysis, data interpretation

VNK: data analysis, data interpretation

SA: data analysis, data interpretation

TMB: data collection, data interpretation

JKB: data collection, data interpretation

SE: data collection, data interpretation

HV: data collection, data interpretation

UV: data collection, data interpretation

AW: data collection, data interpretation

AT: data collection, data interpretation

JW: data collection, data interpretation

HP: data collection, data interpretation

CG: data collection, data interpretation

JKA: data analysis, data interpretation

LF: data collection, data interpretation

LMi: data analysis, data interpretation

BMM: data analysis, data interpretation, funding acquisition

AHM: data collection, data interpretation

FH: data collection

SE: data collection, funding acquisition

CB: data collection, funding acquisition

SW: data collection, funding acquisition

LA: data collection, funding acquisition

KS: data generation, data interpretation

BSW: data generation, data interpretation

SSp: data collection

TEM: data collection, funding acquisition

HLF: data collection, funding acquisition

AC: data collection, funding acquisition

CMH: data collection, funding acquisition

MS: data collection, funding acquisition

JL: data collection, data analysis

KR: data collection, funding acquisition

EG: data collection, funding acquisition

GSK: data collection, funding acquisition

HJG: data collection, funding acquisition

AP: data collection, funding acquisition

MW: data generation, data interpretation, supervision of data analysis

EBB: study design, data interpretation, supervision of data analysis, writing of manuscript

All authors read, edited and approved the final manuscript.

---

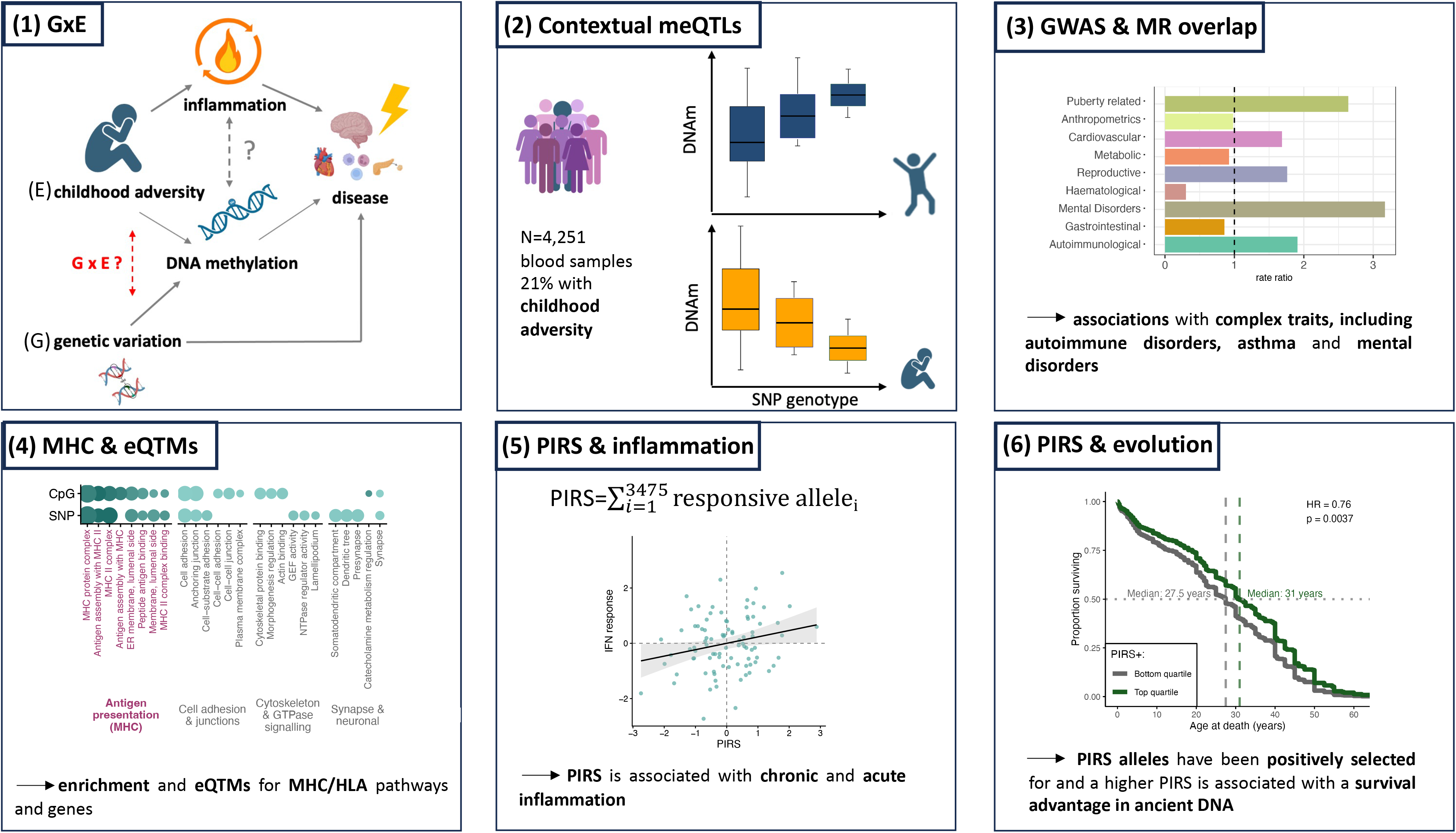

