## Supplemental Material for "Contextual DNA methylation QTLs link adversity responsivity to immune changes and evolutionary trade-offs"

### **Supplementary Methods**

#### 1. Cohorts, phenotypes and omics profiling

ALSPAC cohort

*Sample description*

The Avon Longitudinal Study of Parents and Children (ALSPAC) cohort is a prospective population-based study described in detail in ^1,2^. Pregnant women resident in Avon, UK with expected dates of delivery between 1st April 1991 and 31st December 1992 were invited to take part in the study. 20,248 pregnancies have been identified as being eligible and the initial number of pregnancies enrolled was 14,541. Of the initial pregnancies, there was a total of 14,676 fetuses, resulting in 14,062 live births and 13,988 children who were alive at 1 year of age. When the oldest children were approximately 7 years of age, an attempt was made to bolster the initial sample with eligible cases who had failed to join the study originally. As a result, when considering variables collected from the age of seven onwards (and potentially abstracted from obstetric notes) there are data available for more than the 14,541 pregnancies mentioned above: The number of new pregnancies not in the initial sample (known as Phase I enrolment) that are currently represented in the released data and reflecting enrolment status at the age of 24 is 906, resulting in an additional 913 children being enrolled (456, 262 and 195 recruited during Phases II, III and IV respectively). The phases of enrolment are described in more detail in the cohort profile paper and its update ^1,2^. The total sample size for analyses using any data collected after the age of seven is therefore 15,447 pregnancies, resulting in 15,658 fetuses. Of these 14,901 children were alive at 1 year of age. Of the original 14,541 initial pregnancies, 338 were from a woman who had already enrolled with a previous pregnancy, meaning 14,203 unique mothers were initially enrolled in the study. As a result of the additional phases of recruitment, a further 630 women who did not enroll originally have provided data since their child was 7 years of age. This provides a total of 14,833 unique women (G0 mothers) enrolled in ALSPAC as of September 2021. G0 partners were invited to complete questionnaires by the mothers at the start of the study and they were not formally enrolled at that time. 12,113 G0 partners have been in contact with the study by providing data and/or formally enrolling when this started in 2010. 3,807 G0 partners are currently enrolled. The study website contains details of all the data that is available through a fully searchable data dictionary and variable search tool and at the following webpage: [http://www.bristol.ac.uk/alspac/researchers/our-data](http://www.bristol.ac.uk/alspac/researchers/our-data" \o "http://www.bristol.ac.uk/alspac/researchers/our-data).

Ethical approval for the study was obtained from the ALSPAC Ethics and Law Committee and the Local Research Ethics Committees Informed consent for the use of data collected via questionnaires and clinics was obtained from participants following the recommendations of the ALSPAC Ethics and Law Committee at the time.

A total of 616 participants with available genetic, DNAm and exposure data were included in this study.

*Childhood adversity*

We requested categorical adverse childhood experiences (ACE) scores for exposure between birth and age 16. The construction of these scores has been described in detail previously ^3^. The binary variable CA was coded as exposed (=1) if at least one of the categorial ALSPAC ACE-scores for emotional, sexual or physical abuse was recorded as “exposed”.

*DNA methylation*

We requested normalized and batch corrected blood methylation data for the cohort of 15 years and older, which was assessed on 450K chips. Further information on quality control and normalization is given in [https://github.com/alspac/omics_documentation/tree/main/docs/methylation](https://github.com/alspac/omics_documentation/tree/main/docs/methylation" \o "https://github.com/alspac/omics_documentation/tree/main/docs/methylation). Cell types were estimated based on the Houseman algorithm ^4^.

*Genotyping and imputation*

We requested the imputed genotype data from the cohort of 15 years and older which is based on the Illumina 550 quad chip and imputed using the 1,000 Genomes sample as reference. Details on quality control an imputation is given at [https://proposals.epi.bristol.ac.uk/alspac_omics_data_catalogue.html#orga0e0394](https://proposals.epi.bristol.ac.uk/alspac_omics_data_catalogue.html" \l "orga0e0394" \o "https://proposals.epi.bristol.ac.uk/alspac_omics_data_catalogue.html#orga0e0394).

##### BeCOME cohort

*Sample description*

The Biological Classification of Mental Disorders (BeCOME; trial number on ClinicalTrials.gov: NCT03984084) study as well as inclusion and exclusion criteria are described in detail in ^5^. In brief, the study is conducted at the Max Planck Institute of Psychiatry in Munich, Germany, and includes mainly individuals from the broader Munich area with a broad spectrum of affective, anxiety and stress-related mental disorders and individuals unaffected by mental disorders who responded to diverse study advertisements and were free of psychopharmacological medication for at least the past eight weeks before study participation. All participants underwent in-depth phenotyping procedures and omics assessments on two consecutive days. The study was approved by the local Ethics Committee of the Ludwig Maximilians University (No. 350-14) and written informed consent was obtained from all participants.

At the time of the data freeze, a total of 249 participants with available genetic, DNAm and exposure data was available and could be included in this study.

*Childhood adversity*

For the present study, CA was assessed using the German version of the Childhood Trauma Questionnaire (CTQ) ^6^, a retrospective self-report of exposure to different types of abuse and neglect experiences ^7,8^. If at least one of the subscales (emotional, sexual, physical) met the threshold for moderate abuse, the dichotomous variable CA was coded as exposed.

*DNA methylation*

DNA methylation pre-processing in blood is described in detail in ^9^. Bisulfite-conversion of 400 ng DNA was performed with the EZ-96 DNA Methylation kit (Zymo Research, Irvine, CA). Illumina Infinium MethylationEPIC v1 BeadChip (Illumina, San Diego, CA, USA) was used for epigenome-wide methylation analysis. Methylation data was preprocessed based on an adapted pipeline from ^10^ using the R-package *minfi* ^11^, samples with mean detection p-value > 0.05 and sex mismatches were excluded, normalization was performed based on standard quantile normalization ^12^ and subsequent beta-mixture quantile normalization (BMIQ) ^13^. *Combat* ^14^ was used for batch-correction. *MixupMapper* ^15^ confirmed that no samples mix-up or swaps during the experiment had taken place. Cell types were estimated based on the method proposed by Houseman et al. ^4^.

*Genotyping and imputation*

Genotyping was conducted on Illumina global screening arrays (GSA-24v3-0, Illumina, San Diego, CA, US). SNPs and individuals with a call rate below 98%, a minor allele frequency below 1% or deviation from Hardy-Weinberg-Equilibrium (HWE, *p*-value < 1 × 10^−05^) were excluded. Only unrelated individuals were included and the main multi-dimensional scaling (MDS) components from the IBS matrix were retrieved. Quality control was performed using plink 1.9 (<https://www.cog-genomics.org/plink/>). Imputation was performed using *shapeit2* ^16^ and *impute2* ^17^ based on the 1,000 Genomes Phase III reference sample. Only SNPs with an info-score > 0.6 went into association analysis and were transformed into best-guessed genotypes using a hard-call threshold of 0.9.

*Transcriptomics*

Gene expression levels were assessed using the Lexogen CORALL total RNA-Seq Library Prep Kit, pre-processing and quality control is described in ^18^. Here, we only used the gene-expression of *HLA-DRB5* and *HLA-DRB6.*

##### BioMD-Y cohort

*Sample description*

The biopsychosocial factors of major depression in youth (BioMD-Y) study as well as inclusion and exclusion criteria are described in detail in ^19^. The study comprises children and adolescents with major depressive disorder and their healthy peers.

This study is approved by the Ethics Committee of the Medical Faculty, LMU Munich, REF 297- 09. Participants gave informed consent to participate in the study before taking part.

A total of 551 participants with available genetic, DNAm and exposure data were included in this study. Since the majority of included participants (N=420 out of 551 individuals with available phenotypic, DNAm and genotypic data) were adolescents (14 years or older), this study was included in the combined analysis of adolescents/adults.

*Childhood adversity*

CA was measured including four items presenting emotional, sexual and physical abuse using the Life Event Survey ^20^ and the Munich Event List ^21^. If at least one item was rated with “yes”, the individual was dichotomized into the exposed group.

*DNA methylation*

DNA was extracted according to standard procedures. Bisulfite-conversion of DNA was performed with the EZ-96 DNA Methylation kit (Zymo Research, Irvine, CA). Illumina Infinium MethylationEPIC v1 BeadChip (Illumina, San Diego, CA, USA) was used. Preprocessing of methylation data was performed using an adapted pipeline ^10^ with the *minfi* R package ^11^. Samples with a poor detection p value > 0.05, samples presenting with distribution artefacts in raw beta-values or sex mismatches between estimated sex from methylation data and phenotypic sex were excluded. Normalization of beta-values was performed using stratified quantile normalization ^12^ and subsequently beta-mixture quantile normalization ^13^. Batch effects were sequentially with *Combat* of the *sva* R package ^14^. *MixupMapper* ^15^ confirmed that no samples mix-up or swaps during the experiment had taken place. Cell types were estimated based on the method proposed by Houseman et al. ^4^.

*Genotyping and imputation*

Genotyping was conducted on Illumina global screening arrays (Illumina, San Diego, CA, US). SNPs and individuals with a call rate below 98%, a minor allele frequency below 1% or deviation from HWE (*p*-value < 1 × 10^−05^) were excluded. Only unrelated individuals were included and the main MDS-components from the IBS matrix were retrieved. Quality control was performed using plink 1.9 (<https://www.cog-genomics.org/plink/>). Imputation was performed using *shapeit2* ^16^ and *impute2* ^17^ based on the 1,000 Genomes Phase III reference sample. SNPs with an info-score> 0.6 went into association analysis and were transformed into best-guessed genotypes using a hard-call threshold of 0.9.

##### KORA cohort

*Sample description*

The KORA (Kooperative Gesundheitsforschung in der Region Augsburg) research platform has been collecting clinical and genetic data from the general population in the region of Augsburg, Germany for over 20 years. The KORA FF4 (2013-2014) cohort (n=2,279) is a follow-up study from the KORA S4 (n=4,261) survey carried out 1999-2000. In the baseline examinations all inhabitants of German nationality between the ages of 25 and 74 years were enrolled. Participants completed a lifestyle questionnaire, including details on health status and medication use, underwent standardized examinations with blood samples taken ^22^.

The KORA cohort ethical approval was granted by the ethics committee of the Bavarian Medical Association (REC reference number FF4: #06068) and all were carried out in accordance with the principles of the Declaration of Helsinki. This covers consent for the use of biological material, including genetics. All research participants have signed informed consent prior to taking part in any research activities.

A total of 1,519 participants with available genetic, DNAm and exposure data were included in this study.

*Childhood adversity*

Childhood adversity was based on self-reported questionnaire data (using the questions “Have you ever been hit by family members so hard that you had bruises or scratches?”, “Did you ever have the feeling that family members hated you?”, “Have you ever been sexually abused?”). If at least one of these questions was answered positively (options “a few times”, “often” or “very often”), childhood adversity was set to exposed.

*DNA methylation*

Genomic DNA from 1,928 individuals was bisulfite converted using the EZ-96 DNA Methylation Kit (Zymo Research, Orange, CA, USA) in two separate batches (N=488, N=1,440). Subsequent methylation analysis was performed on an Illumina (San Diego, CA, USA) iScan platform using the Infinium MethylationEPIC v1 BeadChip according to standard protocols provided by Illumina. GenomeStudio software version 2011.1 with Methylation Module version 1.9.0 was used for initial quality control of assay performance and for generation of methylation data export files. Further quality control and preprocessing of the data were performed in R v3.5.1 (https://www.R-project.org/), with the package *minfi* v1.28.3^11^ and following primarily the *CPACOR* pipeline ^23^. Raw intensities were read into R (command read.metharray) and background corrected (bgcorrect.illumina). Probes with detection p-values > 0.01 were set to missing. Before normalization, we removed problematic samples and probes. Forty samples were removed: 2 showed a mismatch between reported sex and that predicted by *minfi*; 33 had median intensity < 50% of the experiment-wide mean, or < 2,000 arbitrary units; and 9 (overlap of 4 with previous) had > 5% missing values on the autosomes. A total of 59,631 probes were removed (some overlapping multiple categories): cross-reactive probes as given in published lists (N=44, 493 ^24,25^); probes with SNPs with minor allele frequency > 5% at the CG position (N=11,370) or the single base extension (N=5,597) as given by *minfi*; and 5,786 with > 5% missing values. A total of 806,228 probes remained for analysis. Quantile normalization was then performed separately on the signal intensities divided into the 6 probe types: type II red, type II green, type I green unmethylated, type I green methylated, type I red unmethylated, type I red methylated ^23^. For the autosomes, QN was performed for all samples together; for the X and Y chromosomes, men and women were processed separately. The transformed intensities were then used to generate methylation beta values, a measure from 0 to 1 indicating the percentage of cells methylated at a given locus. The white blood cell type proportions were estimated using the Houseman algorithm ^4^.

*Genotyping and Imputation*

Genotyping of KORA S4/F4 samples was performed on the Affymetrix Axiom Platform. Genotypes were called with the Affymetrix software and annotated to NCBI build 37. Imputation was performed based on the Haplotype Reference Consortium reference panel (Panel (r1.1), April 2016), using *minimac3* as imputation tool (imputation done on Michigan Imputation server) and with *shapeit2* as a pre-phasing tool.

*Transcriptomics*

After RNA isolation using PAXgene Blood RNA Kit, RNA integrity number (RIN) was measured using the Agilent 2100 Bioanalyzer system. RNA samples with RIN values of approximately 6 or more were selected for mRNA sequencing (poly-A selected). The libraries were prepared using the Illumina stranded mRNA prep ligation kit (Illumina), following the kit's instructions. After a final QC, the libraries were sequenced in a paired-end mode (2x100 bases) in the Novaseq6000 sequencer (Illumina) with a depth of at least 40 million reads per sample. After demultiplexing, FASTQ files from each sample are processed using standard tools ^26^. Alignment to UCSC Genome Browser hg19 human reference genome using *STAR v2.4.2a* ^27^ Unaligned reads are discarded. Sequencing QC was done using *RNASeQC v1.1.8.1* ^28^. Properly aligned reads are then processed with *HTSeq-count v0.6.1* ^29^ to generate read counts which can be interpreted as quantified gene expression. The reads are then normalized for exon length and total sequencing yield to generate Fragments Per Kilobase of transcript per Million mapped reads (FPKM), and this is done through dividing the fragments per gene by the product of length of the gene in kilobase and million reads sequenced.

After sequencing QC, samples QC was done. Samples with < 30 million reads were discarded. Exonic, intronic, itragenic, intergenic and rRNA rates calculated by RNAseQC were examined for outliers but no such outliers were found and no samples were excluded based on these. Only the genes with FPKM of more than 1 in at least 5% of the samples were selected. Number of the selected genes in each sample were calculated. Samples having less than 5,750 genes were excluded. Sex mismatches in the phenotype tables and those discerned from looking at the expression of XIST and UTY genes were also excluded.

##### OPTIMA cohort

*Sample description*

The OPtimized Treatment Identification at the Max Planck Institute of Psychiatry (Munich, Germany) (OPTIMA) study is described in detail in Kopf-Beck et al. ^30^. In brief, Optima is a single-center randomized controlled trial for schema therapy in major depressive disorder among psychiatric inpatients.

A total of 96 participants with available genetic, DNAm and exposure data were included in this study.

*Childhood adversity*

CA was assessed in the same way as in BeCOME, based on the CTQ.

*DNA Methylation*

OPTIMA samples were run together with BeCOME samples on EPIC v1 arrays and quality control was performed in the same way.

*Genotyping and imputation*

OPTIMA samples were run together with BeCOME samples GSA arrays and quality control was performed in the same way.

##### SHIP cohort

*Sample description*

The Study of Health in Pomerania (SHIP) is a population-based project aiming to investigate disease incidences in the northeast of Germany and to analyse the relationship between risk factors, subclinical disorders and disease outcomes ^31^. The SHIP-TREND-0 sample was recruited between 2008 and 2012 and comprises 4,420 people randomly drawn from the adult population of Western Pomerania in Germany.

The study followed the recommendations of the Declaration of Helsinki. The medical ethics committee of the University of Greifswald approved the study protocol, and oral and written informed consents were obtained from each of the study participants.

A total of 440 participants with available genetic, DNAm and exposure data were included in this study.

*Childhood adversity*

CA was assessed via the CTQ ^8^. The same thresholds for exposed as unexposed as in BeCOME were used.

*DNA methylation*

DNA was extracted from blood samples of N=508 SHIP-TREND-0 participants to assess DNAm using the Illumina HumanMethylationEPIC v1 BeadChip array (Illumina, San Diego, CA, USA). Samples were randomly selected based on availability of multiple OMICS data, excluding type II diabetes, and enriched for prevalent myocardial infarction. The samples were taken between 07:00 AM and 04:00 PM, and serum aliquots were prepared for immediate analysis and for storage at -80 °C in the Integrated Research Biobank (Liconic, Liechtenstein). Processing of the DNA samples was performed at the Helmholtz Zentrum München. Preparation and normalization of the array data was performed according to the CPACOR workflow^23^ using the software package R (www.r-project.org). The array idat files were processed using the minfi package. Probes that had a detection p-value above background (sum of per-array methylated and unmethylated intensity values based p-value ≥ 1x10^-16^) were set to missing. Methylation beta values were calculated as proportion of methylated intensity value on the sum of methylated+unmethylated+100 intensities. Arrays with observed methylated+unmethylated+technical problems (±4 SD outside control probe intensity mean) during steps like bisulfite conversion, hybridization or extension, as well as arrays with mismatch between sex of the proband and sex determined by the chr X and Y probe intensities were removed from subsequent analyses. Additionally, only arrays with a call rate ≥ 95% were processed further resulting in N=495 samples with methylation data on 865,859 sites available for subsequent analyses.

To account for potential confounding effects due to blood cell composition, blood cell subtypes were estimated by the Houseman method^4^ and included in the association model.

*Genotyping and Imputation*

A subset of the SHIP-TREND-0 sample was genotyped using the Illumina Human Omni 2.5 array (Illumina, San Diego, CA, USA). Hybridisation of genomic DNA was done in accordance with the manufacturer’s standard recommendations at the Helmholtz Zentrum München. Genotypes were determined using the GenomeStudio 2.0 Genotyping Module (GenCall algorithm). Arrays with a genotyping call rate < 94%, duplicates (based on estimated identical by descent), and mismatches between reported and genotyped sex were removed, leaving N=986 arrays for subsequent analyses. Imputation of genotypes was performed using the HRCv1.1 reference panel and the Eagle and minimac3 software implemented in the Michigan Imputation Server for pre-phasing and imputation, respectively. SNPs with a HWE p-value <  1 × 10^−04^, a call rate < 0.95, or monomorphic SNPs were removed before imputation, as well as SNPs having position mapping problem from genome build b36 to b37, duplicate IDs, or with inconsistent reference site alleles.

##### GLAKU cohort

*Sample description*

The adolescents of the GLAKU (Glycyrrhizin in Licorice) cohort came from an urban community-based cohort comprising 1049 infants born between March and November 1998 in Helsinki, Finland ^32^. In 2009–2011, initial cohort members who had given permission to be contacted and whose addresses were traceable (*N* = 920, 87.7% of the original cohort in 1998) were invited to a follow-up, of which 692 (75.2%) could be contacted by phone (mothers of the adolescents). Of them, 451 (65.2% of those who could be contacted by phone, 49% of the invited) participated in a follow-up at a mean age of 12.3 years (SD = 0.5, range 11.0–13.2 years). The Ethics Committees of the City of Helsinki and the Helsinki and Uusimaa Hospital District approved the study protocol. Written informed consent was obtained from the mother at birth and from parent/guardian and adolescent at the follow-up.

A total of 239 participants with available genetic, DNAm and exposure data were included in this study.

*Childhood adversity*

CA was assessed by the presentation of stressful life events (SLE). Maternal reports were available on SLEs (maternal unemployment, parental separation, and death of a spouse) in follow-ups at ~6 months, ~5 years, ~8 years, and/or ~12 years of age. If the mother reported any SLE in any of the follow-ups, childhood adversity was set to present.

*DNA methylation*

Illumina Infinium HumanMethylation450 BeadChip (Illumina, San Diego, California) was used to determine genome wide DNAm levels. Pre-processing was performed using the R-package *minfi* ^11^. Raw beta-values were normalized using functional normalization ^33^, CpG-sites failing in more than 50% of the samples were removed. Batch-correction was performed using *Combat* ^14^ and non-autosomal CpGs as well as cross-hybridizing CpGs were removed. Cell types were estimated based on the method proposed by Houseman et al. ^4^

*Genotyping and imputation*

DNA was extracted from blood samples (N=80) and saliva samples (N=277) donated at the 2009-2011 follow-up and genotyping was performed with the Illumina OmniExpress Exome 1.2 bead chip at the Tartu University, Estonia in September 2014 according to the standard protocols. Genomic coverage was extended by imputation using the 1000 Genomes Phase I integrated variant set (v3 / April 2012; NCBI build 37 / hg19) as the reference sample and IMPUTE2 software. Before imputing the following QC filters were applied: SNP clustering probability for each genotype > 95%, Call rate > 95% individuals and markers (99% for markers with MAF < 5%), MAF > 1%, HWE p > 1 × 10^−06^. Moreover, heterozygosity, gender check and relatedness checks were performed and any discrepancies removed (N=2). We performed MDS-analysis on the identity by state matrix of quality-controlled genotypes. The first three components depicted the origin admixture and were included as covariates in the regression analyses.

##### Kids2Health

*Sample description*

The Kids2Health study is a Berlin-based case-control study for studying the biological embedding of early-life adversity ^34^. The study population consists of children aged 3-12 years, 294 of them exposed to early-life stress, and 241 unexposed control children. The cohort was also enriched for obese children (BMI >= 95^th^ percentile in 12.3% of participants). Controls were screened to ensure they had not experienced maltreatment, violence, or any severe critical or traumatic life events. General exclusion criteria for all children were severe mental or physical disability, severe chronic diseases, chronic intake of medications affecting the nervous system, severe accidents, and insufficient language skills in German or Arabic. Participants were recruited between 2018 and 2022 from local child welfare and protection services, refugee services, pediatric clinics, the Berlin registry of inhabitants, and online advertisements. Caregivers and children aged six or older gave written informed consent and caregivers received monetary compensation for their participation. The study was approved by the Ludwig-Maximilians-Universität München (LMU) Ethics Committee (18-444).

A subset of 319 non-related children with available genotype and DNAm data at baseline were included in this study.

*Childhood adversity*

Early-life-stress-exposed children in the Kids2Health study were exposed to childhood maltreatment or war- and migration-related traumatic events. Childhood maltreatment was assessed via the Maternal Interview for the Classification of Maltreatment (MICM; ^35^), which was conducted with the caregiver by trained clinicians. Maltreatment severity was coded on a five-point scale using the Maltreatment Classification System (MCS; ^35^), and children were classified as maltreated when meeting the following cutoffs: emotional maltreatment ≥ 2, physical neglect ≥ 2, physical abuse ≥ 1, sexual abuse ≥ 1. War- and migration-related trauma was assessed using the UCLA Trauma Exposure Screener ^36^ and verified by parent report.

*DNA methylation*

DNA was extracted from all available peripheral blood samples at baseline. DNA extraction was conducted in a semi-automated manner using magnetic beads on a Chemagic 360 device (Revvity chemagen Technologie GmbH) using the Chemagic DNA Blood Kit H96. DNA was bisulfite-converted using the EZ-96 DNA Methylation Kit (Zymo Research Corperation) and then run on the EPIC v2.0 Bead Chip (Illumina Inc., San Diego, USA). Assignment of samples to plates and chips was randomized for age, reported sex, early-life stress status, subproject, and time point. Methylation data were pre-processed in R according to a customized *minfi* ^11^ workflow (<https://github.molgen.mpg.de/mpip/EPIC_Preprocessing_Pipeline>). Data were normalized via stratified quantile normalization ^12^ followed by beta-mixture quantile normalization (BMIQ) ^13^ and batch-corrected for plate, slide and array using *Combat* via the *sva* R package ^14^. We excluded probes on chromosomes X and Y, probes with mapping inaccuracies and flagged probes as reported by Illumina, and probes with a detection p-value > 0.01. For replicate probes, only the probe with the lowest detection p-value was retained. Samples were excluded in case of mean detection p-value > 0.05, mismatches of epigenetically predicted and phenotypic sex, or mismatches of genotypic and methylation data identified with *MixupMapp*er ^15^. Cell type composition of the peripheral blood samples was estimated with the R package *epidish* ^37^ using a reference panel of 12 leukocyte cell types ^38^.

*Genotyping and Imputation*

Genotyping was conducted on Illumina global screening arrays (Illumina, San Diego, CA, US). SNPs and individuals with a call rate below 98%, a minor allele frequency below 1% or deviation from HWE (*p*-value < 1 × 10^−05^) were excluded. Only unrelated individuals were included and MDS-components were retrieved. Quality control was performed using plink 1.9 (<https://www.cog-genomics.org/plink/>). Imputation was performed using *shapeit2* ^16^ and *impute2* ^17^ based on the 1,000 Genomes Phase III reference sample. SNPs with an info-score> 0.6 went into association analysis and were transformed into best-guessed genotypes using a hard-call threshold of 0.9.

##### LISA south cohort

*Sample description*

The influence of Life-style factors on the development of the Immune System and Allergies in East and West Germany (LISA) study is a population-based birth cohort study. A total of 3,094 healthy, full-term neonates were recruited between 1997 and 1999 in Munich, Leipzig, Wesel and Bad Honnef. The participants were not pre-selected based on family history of allergic diseases. Detailed descriptions of the LISA study have been published elsewhere ^39^. Blood was collected at the age of 6 and 10 years. Approval was given by the local Ethics Committees and written consent from participant’s families was obtained. A total of 222 participants with available genetic, DNAm and exposure data were included in this study.

*Childhood adversity*

CA events were defined as exposure to at least one of the following events up to the age of 6 years: divorce, severe disease, death, unemployment. An aggregated binary variable was constructed based on these events, resulting in 113 (50.9%) participants experiencing CA events and 109 (49.1%) without any CA event until the age of 6.

*DNA methylation*

DNAm was measured in blood using the MethylationEPIC v1 BeadChip (Illumina, Inc., San Diego, CA). Cell type proportions were estimated with the Houseman method ^4^. Details are described elsewhere ^40^.

*Genotyping and imputation*

588 individuals from the LISA study were analysed using the Affymetrix Human SNP Array 5.0 and 88 individuals were analysed using Affymetrix Human SNP Array 6.0. Genotypes were called using BRLMM-P algorithm (5.0), respectively BIRDSEED V2 algorithm (6.0). In each of the two data sets, criteria for exclusion of individuals were: a call rate below 95%, heterozygosity outside mean +/- 4sd, a failure of the sex check or a failure of the similarity quality control using MDS analysis based on IBS. Criteria for exclusion of variants were: a call rate below 95%, a MAF < 0.01 and a HWE p-value < 1 × 10^−05^. Haplotype phasing and imputation in the HRC panel version1.1 was performed using the University of Michigan server ^41^. Variants with an imputation quality score of r^2^ < 0.5 or out of HWE p < 1x10^-12^ were excluded.

##### UK Biobank

*Sample description*

UK Biobank is a population-based cohort of 500,000 participants aged 40-69 years, recruited between 2006 and 2010. All participants provided informed consent. Further information on the cohorts and how to apply for access is available in ^42^ and online (<https://community.ukbiobank.ac.uk/hc/en-gb>).

*Childhood adversity*

Childhood adversity was defined based on the traumatic events questionnaire. If any of the questions: “Physically abused by family as a child”, “Felt hated by family member as a child” or “Sexually molested as a child” was ticked as “yes”, CA was coded as 1, otherwise as 0. *Genotyping and imputation*

Pre-processed and qc-ed imputed genotypes as well as genetic PCs were downloaded from the UKB-RAP. Further information on genotyping, quality control and imputation is available online (<https://biobank.ndph.ox.ac.uk/ukb/label.cgi?id=263>) as well as fully described in ^42^. *Plasma protein levels*

Pre-processed and quality controlled normalized protein expression values of plasma protein levels were downloaded from UKB-RAP. The assessed plasma proteomics using the OLINK technology are described in detail in ^43^.

##### E-Risk

*Sample description*

Participants were members of the Environmental Risk (E-Risk) Longitudinal Twin Study, which tracks the development of a nationally representative birth cohort of 2,232 British twin children. The sample was drawn from a larger birth register of twins born in England and Wales in 1994-1995 ^44^. Full details about the sample are reported elsewhere ^45^. Briefly, the E-Risk sample was constructed in 1999-2000, when 1,116 families (93% of those eligible) with same-sex 5-year-old twins participated in home-visit assessments. This sample comprised 56% monozygotic (MZ) and 44% dizygotic (DZ) twin pairs; sex was evenly distributed within zygosity (49% male). Families were recruited to represent the UK population of families with newborns in the 1990s, on the basis of residential location throughout England and Wales and mother’s age.

Follow-up home-visits were conducted when children were aged 7, 10, 12 and 18 years (participation rates were 98%, 96%, 96% and 93%, respectively). Home-visits at ages 5, 7, 10, and 12 years included assessments with participants as well as their mother (or primary caregiver); the home-visit at age 18 included interviews only with the participants. Each participant in a twin pair was assessed by a different interviewer. There were 2,066 E-Risk participants who were assessed at age 18. The average age of the participants at the time of the assessment was 18.4 years (SD = 0.36); all interviews were conducted after the 18th birthday.

The Joint South London and Maudsley and the Institute of Psychiatry Research Ethics Committee approved each phase of the study (NRES 1997/122). Parents gave informed consent and twins gave assent between 5-12 years and then informed consent at age 18.

*Childhood adversity*

Adolescent experiences of stress and violence between 12 and 18 years of age were assessed at age 18 using the Juvenile Vicitimization Questionnare (JVQ-R2), adapted as a clinical interview, details are given in ^46,47^. In brief, seven domains of adverse experiences (crime victimization, peer or sibling victimization, cyber bullying, sexual abuse, physical, maltreatment, family violence, and neglect) were assessed.

*Genotypes and imputation*

Genotyping was performed using Illumina HumanOmni Express arrays as described in ^48^. After quality control, additional SNPs were imputed using *impute2 ^17^* and the 1000 Genomes reference dataset ^49^.

*Inflammation markers*

Plasma samples were available for 1,448 participants (see ^46^ for details). Briefly, high-sensitivity CRP was measured using enzyme-linked immunosorbent assay (ELISA) (Quantikine ELISA Kit DCRP00, R&D Systems) following the manufacturer’s protocol. Plasma IL-6 levels were measured using ELISA (Quantikine HS ELISA Kit HS600C, R&D Systems) following the manufacturer’s protocol. Plasma suPAR levels were analyzed using ELISA (suPARnostic AUTO Flex ELISA, ViroGates A/S) following the manufacturer’s protocol. Due to skewness, CPR and IL6-values were log-transformed. For all three markers, outliers deviating more than 4 SD from the respective marker’s mean were excluded.

#### 2. Statistical analysis

##### Meta-analysis across adult/adolescent cohorts

In each cohort we tested for contextual methylation quantitative trait loci (contmeQTLs) using linear regression models with the specific CpG-site as outcome and the interaction of CA and the specific SNP-genotype (coded as 0-1-2) as predictor, covarying for important confounders (age, sex, genetic PCs, estimated cell types, CpG cg05575921 indicative of smoking status ^50^). Only combinations including meQTLs identified in GoDMC ^51^, i.e. 66,577,913 significant *cis* meQTLs (at most 1MB distance between CpG-site and SNP, made up of 6,316,998 unique SNPs and 183,207 unique CpG-sites, p< 1x10^-08^) and 3,657,300 significant *trans* meQTLs (more than 1MB distance between CpG-site and SNP as well SNPs located on a different chromosome than the CpG-site, made up of unique 893,991 SNPs and unique 19,147 CpGs, p<1x10^-14^), were tested. We combined results across cohorts using random-effects meta-analysis based on the DerSimonian-Laird random effects model as implemented in an extension of METAL ^52^. To assure that results were not driven by one specific cohort, we restricted the meta-analysis to combinations of SNPs and CpGs which were available in at least three of the six cohorts. If several contmeQTLs for the same CpG-site were present, the contmeQTL SNP with the lowest interaction p-value was chosen. Afterwards, we applied multiple testing correction using false-discovery-rate (FDR) with a threshold of 0.05.

##### Sex-specific results

We checked post-hoc if any of the 5,090 identified *cis* and 30 identified *trans* contmeQTLs showed sex-specific effects. For this, we used the four cohorts where individual level data was available to us (ALSPAC, BeCOME, BioMD-Y and OPTIMA), stratified each cohort by genetic sex and reran the interaction analysis. Afterwards, we compared if effect directions where different between males and females. No contmeQTL showed significant differences between females and males which were consistent across the cohorts.

##### Celltype specific effects

To evaluate if any of the 5,120 CpG-sites regulated by contmeQTLs were cell-type specifically methylated, we checked if they overlapped with the top 250 and the top 1,000 marker CpG-sites for blood cells (B-cells, T-cells, granulocytes, natural killer cells, monocytes and tissue-resident macrophages) identified in ^53^.

##### Sensitivity analyses: including BMI as covariate and running the fully saturated model

The primary model included the same covariates as the meQTL analysis on which our SNP-CpG pairs were based ^51^: sex, age, genetic principal components, celltype proportions and cg05575921 as methylation-based smoking proxy. BMI was not included in the primary model because it may be partly a consequence of the exposure. In a sensitivity analysis, we additionally included BMI, because BMI is associated with both CA and DNAm ^54,55^. Since covariates entered only as main effects do not control for their confounding of a gene-by-environment interaction, we also added the interactions of every covariate with genotype and with childhood adversity ^56^. We ran these models for the significant 5,120 contmeQTLs and in the two largest cohorts with individual genotype, methylation and phenotypic data available to us (n=616 in ALSPAC, n=551 in BioMD-Y). BMI was only available for a small subset of participants (n=78) in BioMD-Y, therefore we did not include BMI but only the interactions of the original covariates with genotype and with CA in the model. In ALSPAC, BMI was available for 571 participants. Here, we included BMI as covariate and ran the fully saturated model in this subset.

##### Overlap with variance methylation quantitative trait loci (vmeQTLs)

To check for overlap with vmeQTLs, we used the data of Ek et al. ^57^ who identified 7,195 significant CpGs regulated by vmeQTLs in blood. Overall, 5,115 of these CpGs were available in our meta-analysis and 411 overlapped with significant contmeQTLs. To evaluate if this overlap was statistically significant, we randomly selected 5,115 CpGs which had been tested in the meta-analysis and repeated this 1,000 times. For each run, we checked how many of the random CpGs overlapped with CpGs regulated by vmeQTLs and if this overlap was greater than 411. Based on these counts, an empirical p-value was derived. We followed the same logic for the enrichment of SNPs: of the 6,456 unique SNPs from Ek et al. ^57^, 5,466 SNPs were available in our meta-analysis and 43 overlapped with SNPs implicated in significant contmeQTLs. We randomly selected subsets of 5,466 SNPs which had been tested in the meta-analysis 1,000 times and for each run checked if the overlap with vmeQTLs was greater than 43.

##### Enrichment for chromatin states (chromHMM marks)

We used the histone ChIP-seq peaks from the Roadmap Epigenomics project, i.e. the pre-processed consolidated broad peaks from the uniform processing pipeline of the Roadmap project (<http://egg2.wustl.edu/roadmap/data/byFileType/peaks/consolidated/broadPeak/>). These were annotated to chromatin-state signatures (chromHMM) using hidden Markov models ^58^. Overlapping SNP-positions and CpG-position with the 15 chromHMM states were evaluated for all SNPs and CpGs included in the meta-analyses as well as for SNPs and CpGs included in significant contmeQTLs. Enrichment for chromHMM states was tested using Fisher-tests for each chromHMM state individually, p-values were Bonferroni-corrected over 15 states and 2 marker types (SNP and CpG).

##### Enrichment for GO-terms

SNPs and CpGs were first assigned to genes based on closest distance using *matchGenes* in the R-package *bumphunter* ^59^. Matched genes from all tested CpGs (n=18,235 unique genes) and all tested SNPs (n=17,105 unique genes) were used as background sets and tested against the 3,265 unique genes mapping to contmeQTL CpGs and the 3,284 unique genes mapping to contmeQTL SNPs. Pathway enrichment for GO-terms was conducted using FUMA ^60^ (<https://fuma.ctglab.nl>). P-values from FUMA were FDR-adjusted for the number of terms tested within the specific GO-set (biological processes, cellular components and molecular function) and further Bonferroni-corrected for all three tested sets.

Expression quantitative trait methylation (eQTM) analysis in KORA and BeCOME

In KORA, we utilized *MatrixEQTL* ^61^ version 2.3 to evaluate linear associations between gene expression and DNA methylation (eQTM) on the 5,115 CpGs regulated by contextual meQTLs, either in *cis* or *trans*. The 1,519 samples with available information for CA from KORA FF4 were included in the analysis. To account for technical variation, both methylation and expression data were adjusted for technical covariates. Specifically, the methylation data was regressed on the top 10 PCs, while the expression data was regressed on a selected subset of principal components (PC 1, 3, and 4) which capture significant sources of technical variation. Association results were corrected for age, sex, smoking and estimated cell counts and corrected for multiple testing based on FDR of 0.05. We only considered cis-acting eQTMs with a window size of 500 kb. In BeCOME, we ran eQTM for the two HLA-genes which presented with significant eQTMs in KORA, *HLA-DRB5* and *HLA-DRB6.* Associations were covaried for genetic PCs, estimated celltypes, age, sex, and smoking, based on the *AHRR*-CpG cg05575921.

##### Enrichment for genome-wide association studies (GWAS)

Genome-wide significant GWAS associations (p<5.2x10^-08^) were extracted from the MRC IEU OpenGWAS platform (<https://gwas.mrcieu.ac.uk>) following the same strategy as described previously ^62^: we focused on phenotypes originating from the batches UK Biobank study, the NHGRI-EBI GWAS Catalog, and brain imaging phenotypes based on UK Biobank data. This list was filtered for duplicates and high similarity in trait names resulting in a list of 7,503 traits and 11,197,180 individual association results. These results were subsetted to all SNPs included in the contmeQTL meta-analysis resulting in 133,121 unique SNPs, associated with 1,975 unique study traits. Associated traits were categorized into different domains. Domains were created using ChatGPT 3.5 (<https://chatgpt.com>) and the prompt “How would you group these traits into different biological or disease-associated domains?” and manually curated afterwards, resulting in 35 unique trait domains (see **Table S3**). Afterwards, this list was filtered for SNPs involved in significant contmeQTLs, resulting in 2,307 unique SNPs and 33 unique trait domains. For each trait domain, we tested if the rate of GWAS associations for SNPs acting as contmeQTLs was higher as compared to the rate of GWAS associations for SNPs not acting as contmeQTLs using Poisson rate tests for enrichment, followed by FDR-correction. Domains with less than 1% frequency in the contmeQTL CpGs were removed. Domains with less than 1% frequency in the contmeQTL SNPs and not present in the MR-analysis were removed.

##### Enrichment for epigenome-wide association studies (EWAS)

Results from the EWAS catalogue were downloaded from <https://www.ewascatalog.org/download/> on Feb 19^th^ 2024 and filtered for CpG-sites epigenome-wide significantly associated (p<9x10^-08^) with at least one EWAS trait resulting in 1,257,086 individual association results. We filtered these results for all CpG-sites included in the meta-analysis, resulting in 146,053 unique CpG-sites, associated with 1,990 unique study traits. Associated traits were categorized into different domains. Domains were created using ChatGPT 3.5 (<https://chatgpt.com>) and the prompt “How would you group these traits into different biological or disease-associated domains?” and manually curated afterwards, resulting in 39 unique trait domains (see **Table S5**). Afterwards, this list was subsetted to CpGs involved in significant contmeQTLs, resulting in 4,815 unique CpGs and 32 unique trait domains. For each trait domain, we tested if the rate of EWAS associations for CpGs regulated by contmeQTLs was higher as compared to the rate of EWAS associations for CpGs not regulated by contmeQTLs using Poisson rate tests for enrichment, followed by FDR-correction. Domains with less than 1% frequency in the contmeQTL CpGs were removed.

Overlap with Mendelian Randomization hits

To evaluate if the identified CpGs regulated by contmeQTLs could be potentially also causally involved in complex traits, we used the Mendelian randomization (MR)-results from Richardson et al. ^63^ who ran a systematic MR analysis using blood meQTL SNPs as instruments and complex traits as outcome variables. Traits were manually mapped to trait domains (see **Table S7**). For each trait domain, we tested if the rate of MR associations for CpGs regulated by contmeQTLs was higher as compared to the rate of MR associations for CpGs not regulated by contmeQTLs using Poisson rate tests for enrichment. Domains with less than 1% frequency in the contmeQTL MR CpGs were removed.

Overlap with glucocorticoid receptor response meQTL

Knauer-Arloth et al. ^64^ identified glucocorticoid receptor (GR)-response meQTLs, i.e. SNPs associated with DNAm changes after stimulation with dexamethasone. We overlapped their list of 104,828 significant GR-response meQTLs (before LD-clumping) with our significant contmeQTLs. This resulted in only 7 combinations with the same CpG-site and the same SNP in both lists, indicating no significant overlap.

##### Construction of polygenic interaction responsivity score

For the construction of a polygenic interaction responsivity score (PIRS), we used all SNPs identified as contmeQTLs. To retain a set of independent SNPs, we first performed LD-clumping as implemented in the R-package *ieugwasr* based on the interaction p-values in the initial meta-analysis, a window-size of 1 MB and r^2^ of 0.2. If a SNP was interacting with CA on more than one CpG-site, the combination with the minimal p-value was kept. This resulted in 3,475 independent SNPs. Second, for the construction of the PIRS, we focused on those initial cohorts where individual level methylation and genotypes data was available to us (i.e., ALSPAC, BeCOME, BioMD-Y, OPTIMA). Specifically, to identify the responsive allele for each SNP*_i_* (i=1,..,3475) in each cohort*_j_* (j=1,..,4), for each SNP-allele A1 and A2, we computed the absolute mean DNAm change |ΔA1_ij_| and |ΔA2_ij_| between individuals with and without exposure to CA. To account for homozygosity, individuals with two copies of the respective allele counted twice, whereas heterozygous individuals contributed once. Finally, for each SNP*_i_*, we computed ΔA1*_i_* and ΔA2*_i_* across all cohorts*_j_* as weighted mean of the individual cohorts’ |ΔA1_ij_| and |ΔA2_ij_|. As cohort-specific weights *w_j_* we chose the cohort specific sample sizes relative to the overall sample size across all cohorts assuring that all weights summed up to 1. If ΔA1_i_ > ΔA2_i_, A1 was set as responsive allele for SNP_i_, otherwise A2. The final PIRS was based on the sum of copies of the responsive alleles (see Table **S13**) and was calculated using plink 1.9 (<https://www.cog-genomics.org/plink/>).

##### Association of PIRS with inflammatory markers in UK Biobank

All SNPs included in the PIRS were available in UK Biobank. We computed the PIRS as the average of the sum of responsive alleles in the UK Biobank cohort, divided by the number of available PIRS SNP alleles to account for the number of available PIRS SNPs per individual, and associated it with plasma protein levels of pro-inflammatory markers. Markers in the OLINK inflammation panel missing in more than 20% of individuals (n=0) as well as individuals missing more than 20% of markers (n=1,996) were removed. Single missing markers levels were imputed based on median imputation. For analyses, we focused on the 31 pro-inflammatory markers included in the panel. These selected proteins comprise core pro-inflammatory cytokines, TNF superfamily members, IL-17 family cytokines, and multiple CC- and CXC-chemokines as well as alarmins, all of which are well-established mediators of innate and adaptive inflammatory responses ^65-69^, namely CCL11, CCL13, CCL19, CCL2, CCL20, CCL23, CCL25, CCL3, CCL4, CCL7, CCL8, CD40, CXCL1, CXCL10, CXCL11, CXCL5, CXCL6, CXCL8, CXCL9, IFNG, IL12B, IL17A, IL17C, IL1A, IL6, IL18, LTA, OSM, S100A12, TNF, TNFSF14.

To test the association of the PIRS with the inflammatory marker profile, the 31 z-scored markers were modelled jointly in a multivariate linear regression with scaled PIRS and CA status as predictors, adjusted for sex, age, BMI and the first five genetic principal components. The joint null hypothesis that the marker-specific coefficients of the PIRS are all zero was tested with a Wald test, with 31 degrees of freedom, using the heteroskedasticity-consistent sandwich covariance matrix of the stacked coefficients (R package *sandwich*). To assess the effect of the PIRS within the CA and no CA groups, we ran single marker associations in each group separately (CA: N=3,472 and no CA: N=17,371) based on linear regressions of the PIRS as predictor and each pro-inflammatory marker as dependent variable, correcting for genetic sex, age, BMI and the first five genetic PCs. We computed the test-statistic of a paired Wilcoxon rank sum test, based on the standardized regression coefficients of the PIRS for all 31 markers in the CA and no CA groups. Afterwards, we ran 10,000 permutations, randomly shuffling the CA and no CA status (together with the covariates) and recomputing the effect sizes in each group and the test statistic of the paired Wilcoxon rank sum test for each permutation. The empirical p-value was then computed as ratio of test-statistics equal to or more extreme than the original test-statistic and the number of permutations.

##### Association of PIRS with inflammatory markers in E-Risk

In the E-Risk cohort, 2,228 PIRS SNPs were available. For the remaining SNPs, we searched for proxies using the R-package *LDlinkR* ^70^ and included a SNP as proxy if it presented with at least R^2^=0.8. The final PIRS included 3,106 SNPs, i.e. 90% of the original score SNPs. We computed the PIRS as the sum of responsive alleles and divided it by the number of available PIRS SNP alleles to account for the number of available PIRS SNPs. We restricted the analysis to DZ twin pairs as our main objective was to examine the association between individual differences in the PIRS and inflammation while accounting for the familial dependence between twins. DZ twins share, on average, approximately 50% of their segregating genetic variants and therefore exhibit meaningful within-family variation in the PIRS. We assessed the association of the PIRS with CRP (available in n=625 DZ twins, 216 with CA), IL-6 (available in n=626 DZ twins, 219 with CA) and suPAR (available in n=628, 219 with CA) using linear mixed models in the *lme4* R-package with the family-ID as random effect, genetic sex, the first 4 genetic PCs as well as BMI at age 18 were included as covariates. All twins were aged 18 years. We assessed cluster-robust variances via sandwich estimators using family-ID as clustering variable.

##### Association of PIRS with viral infection in PBMCs

To test whether contmeQTLs predicted individual differences in immune response directly, we applied the PIRS to an experiment examining how genetics and transcriptomics relate to immune response by Randolph et al. ^71^. This study generated imputed genotypes for 89 samples with paired since-cell RNA-sequencing on blood samples exposed to a mock condition or the influenza A virus. For each genetic variants within the PIRS, we extracted dosages from the published genotype matrix by matching on chromosome, base-pair position, and alleles (using the liftOver tool to get GRCh38 coordinates). Similar to the UK Biobank calculation, the PIRS was computed as the sum of responsive allele dosages across SNPs (N = 2,876 PIRS variants were available in this cohort, i.e. 83% of all PIRS variants). To account for population stratification, we computed genotype principal components (PCs) using common SNPs (MAF > 5%). First, we filtered based on LD clumping (r² < 0.2, window of 200 Kb) using the *bigsnpr* package ^72^. The first 5 PCs were computed from the LD-pruned SNP set via randomized singular value decomposition as implemented in *bigstatsr*.

Normalized pseudobulk expression profiles for five immune cell types (monocytes, CD4+ T cells, CD8+ T cells, NK cells, and B cells) from Randolph et al. ^71^ were used to calculate scores measuring transcriptional response to infection (IFN response score). We computed the IFN response score as the gene expression for genes belonging to the MSigDB ^73,74^ Hallmark IFN-alpha and IFN-gamma gene sets. Gene expression was Z-scored across individuals within each condition and averaged; the response score was calculated as the flu minus mock condition expression, so that higher scores indicate stronger transcriptional response to infection. We calculated the IFN transcriptional response score as a peudobulk score, using the average transcriptional response across all cell-types for each sample. The transcriptional response score and the PIRS were adjusted for age, age^2^, sex, the first five genetic PCs, ancestral background, and cell-type proportions and standardized (mean = 0, and SD = 1) prior to analysis. Associations between the PIRS and transcriptional response scores were tested by correlations between the PIRS and the transcriptomic score. Furthermore, we computed cell-type specific scores in the same manner, adjusting for age, age^2^, sex, the first five genetic PCs and ancestral background.

##### Evolutionary analyses on PIRS

###### Directional selection contmeQTLs

To understand the evolutionary history of contmeQTLs, we looked for evidence of recent positive selection using a large dataset of genome-wide genotypes and selection estimates from a recent study on > 15,000 humans from the past ~20,000 years ^75^. First, we compared the rate of genome-wide significantly positively selected LD-independent contmeQTLs (the same variants used in the PIRS), calculated as the proportion of positively selected PIRS alleles (responsive allele selection > 0 and FDR adjusted p-value < 0.05). Given that we are starting from a functional set of variants (meQTLs with context-dependent effects), we tested whether this ratio was higher than expected compared to similar functional variants (meQTLs that did not show context-dependent effects). To do this, we compared the rate of positively selected PIRS SNPs to random samples of matched, non-contextual (control) meQTLs. These control meQTLs were LD-pruned using the same parameters as in the contmeQTL pruning, and were at least 1 MB away from any contmeQTL. We generated 10,000 random samples of LD-pruned non-contmeQTLs, matched to have the same number of SNPs and similar allele frequency distribution (using 5% bins) as the PIRS pool. For each permutation, we computed the proportion of significantly positively selected non-contmeQTLs (selection > 0 and FDR adjusted p-value < 0.05). Empirical p-values were calculated as the proportion of null-permutation positive selection rates greater than or equal to the observed positively selected PIRS rate. As a sensitivity analysis, we additionally fit a Firth-penalized logistic regression (in order to better account for the rare positive-selection outcome than a standard model) predicting positive-selection status from contmeQTL status (contmeQTL vs. non-contextual meQTL) and included allele frequency as a covariate. Since both the contmeQTL and background meQTL sets are LD-pruned using identical clumping parameters, variants were treated as independent observations.

###### PIRS prediction of estimated age-at-death in ancient humans

To test whether contmeQTLs may be associated with better survival in ancient humans, we tested whether PIRS predicted estimated age-at-death in N=983 ancient individuals with genotype data using the Allan Ancient DNA Resource, with imputed genotypes and estimated age-at-death ^75,76^. We restricted PIRS construction to contmeQTLs whose responsive allele showed significant positive selection in a recent genome-wide scan (selection > 0 and FDR < 0.05, N=18 PIRS SNPs) ^75^, summing responsive allele dosages across these LD-independent variants to compute the PIRS+ for each individual.

For each individual, we also gathered sample date (years before present), genetic sex, and 1240k-panel autosomal target SNP coverage (included as a data quality covariate). The score was standardized (mean=0, SD=1) and residualized on sample age to remove temporal trends. Since all ancient individuals are, by definition, deceased, we modeled age-at-death as a survival time using Cox proportional-hazards regression (R-package *survival*), predicting age-at-death from the PIRS+ and adjusting for sample age, genetic sex, and genotype target coverage. To assess the robustness of the observed associations, we fit two sets of models: one with the continuous PIRS+ across all samples and another comparing the top versus bottom PIRS+ quartiles.

Next, to determine whether this survival association reflected contextual meQTL effects specifically, rather than a generic property of any positively-selected variant set, we repeated the analysis across 10,000 matched-null permutations using positively-selected non-contextual meQTLs (meQTL+). The null distribution was built from a genome-wide background pool of LD-pruned, non-contextual meQTLs at least 1 MB away from any contmeQTL, and under significant positive selection (selection > 0, FDR < 0.05, N=105 independent variants). For each permutation, we drew, without replacement, a random control set of positively selected non-contextual meQTLs matched to the number of positive-selection PIRS variants (N=18), stratified by minor-allele-frequency (MAF) decile to match the MAF distribution of the FDR-significant background pool. Permuted scores were computed as the sum of dosages for the sampled positively selected non-contextual meQTLs, residualized on sample age, and used in the same survival analysis described above to generate a distribution of effect sizes. Empirical p-values were calculated as the proportion of null-permutation Hazard Ratios less than or equal to the observed positively selected PIRS Hazard Ratio.

###### Gene set enrichment near selected contmeQTLs

To characterize the biological pathways influenced by positive selection at contmeQTLs, we mapped each LD-independent contmeQTL (PIRS SNPs) to nearby protein-coding genes (±500 kb based on hg19 coordinates) and performed genome-wide over-representation testing for Gene Ontology (GO) ^77,78^, KEGG ^79^, and REACTOME ^80^ terms (using the R-package *gprofiler2* ^81^). We tested three gene sets separately: genes near positively-selected contmeQTLs (responsive allele selection > 0 and FDR adjust p-value < 0.05) that do not also map to a negatively-selected contmeQTL, genes near negatively-selected contmeQTLs (responsive allele selection < 0, FDR adjusted p-value < 0.05) that do not also map to a positively-selected contmeQTL, and genes near any significantly selected non-contextual meQTL (regardless of selection direction), used as a comparison set to gauge the specificity of the top positively-selected contmeQTL terms.

###### Genetic risk for complex traits

We next tested whether positively-selected contmeQTLs (PIRS responsive allele selection statistic > 0, and selection FDR < 0.05) are associated with increased genetic risk signal for autoimmune and neuropsychiatric traits compared to matched, positively-selected non-contextual (control) meQTLs. Autoimmune/inflammatory traits included asthma ^82^ and type 1 diabetes ^83^. Neuropsychiatric traits comprised six genomic factors from a recent comprehensive genomic structural equation model of psychiatric traits ^84^ (Compulsive Disorders [F1], Schizophrenia-Bipolar [F2], Neurodevelopmental [F3], Internalizing [F4], Substance Use [F5], and a general psychopathology P-factor). A hair color GWAS ^85^ (red hair, GWAS catalogue accession number: GCST90041837) was chosen as negative controls as being very unlikely related to immune function. Variants were filtered per-GWAS to have MAF > 1%, and, where available, imputation INFO ≥0.6. For each trait, we computed the median GWAS Z-score of positively-selected PIRS (matched to the responsive allele) and compared it to a null distribution built from the same positively-selected control meQTLs described above (N=105), allele-frequency-stratified to match the allele frequency distribution of the observed contmeQTL set and matched on N. For each of 10,000 permutations, we computed the median GWAS Z-score of the matched sample of positively-selected non-contextual meQTLs per trait. Empirical p-values were calculated as the proportion of null-permutation median GWAS Z-scores greater than or equal to the observed positively selected PIRS median GWAS Z-score.

###### Sensitivity tests for the evolutionary analyses

In the main analyses, all evolutionary analyses of contmeQTLs used a strict genome-wide selection cutoff (FDR < 0.05) from the selection scan of Akbari et al. To test whether our evolutionary findings were robust across selection thresholds, we repeated all selection threshold-dependent analyses across 14 selection p-value cutoffs (1×10^‑08^, 5×10^‑08^, 1×10^‑07^, 5×10^‑07^, 1× 10^‑06^, 5×10^‑06^, 1×10^‑05^, 5×10^‑05^, 1×10^‑04^, 5×10^‑04^, 1×10^‑03^, 5×10^‑03^, 0.01, and 0.05). For context, positively selected contmeQTLs with an FDR < 0.05 corresponds to a selection p-value between 5×10^-07^ and 1×10^-07^. At each cutoff, a variant was considered positively selected if its selection statistic was positive and its selection p-value fell below the cutoff, and negatively selected if its selection statistic was negative and its selection p-value fell below the cutoff. The rule was applied to both contmeQTLs (with selection direction aligned to the responsive allele) and to non-contmeQTL meQTLs (non-contmeQTLs), so that the selected variant sets were redefined at every cutoff and comparable. All statistical tests were repeated across each selection p-value threshold. All analyses used the LD-pruned set of contmeQTLs (the same set used in the PIRS) and an LD-pruned set of non-contmeQTLs (LD-pruned using the same parameters, and at least 1MB away from any contmeQTL), in the following sections all references to “contmeQTLs” refers to the set of variants used in the PIRS analysis. All downstream permutation analyses used 10,000 permutations.

Enrichment for positive selection

At each cutoff, we tested whether contmeQTLs were more likely to be positively selected than non-contmeQTLs using Firth logistic regression (in order to better account for rare selection events), with positive selection as outcome and contmeQTL status and allele frequency as predictors. We complemented this with a permutation test, in which we repeatedly drew sets of non-contmeQTLs of the same size as the contmeQTL set, matched on allele frequency (based on bins of 5%), and recorded the proportion of positively selected variants in each set. The empirical p-value was the proportion of permuted sets in which this proportion was equal to or larger than that observed for contmeQTLs.

We used these selection enrichment analyses to guide all downstream evolutionary analyses. For the survival, GWAS, and pathway analyses, we restricted these to only examine contmeQTLs at selection p-value cutoffs ≤ 1×10^‑04^, because these were the cutoffs at which both the Firth regression (OR significantly > 1) and the permutation test (empirical p < 0.05) showed significant enrichment of contmeQTLs for positive selection relative to non-contmeQTLs.

Survival in ancient individuals

At each selection p-value cutoff, we recomputed the PIRS+ from the ancient DNA genotypes. The PIRS+ was calculated as the sum of responsive alleles across all contmeQTLs positively selected at that selection p-value cutoff, so that the number of included SNPs varied with the threshold. The score was standardized after being residualized on sample age (thousands of years before present) and divided into quartiles. As in the main analysis, we compared individuals in the top and bottom quartiles using Cox proportional-hazards regression of estimated age-at-death, covarying for sample age, molecular sex and the number of autosomal 1240k target SNPs covered per sample. To obtain a null distribution, we drew 10,000 random sets of LD-pruned non-contmeQTLs positively selected at the same selection p-value cutoff, of the same size as the PIRS+ set and stratified by allele frequency decile, computed a score from each set in the same way and fitted the same Cox model. The empirical p-value was the proportion of permuted hazard ratios (HRs) equal to or lower than the observed HR.

Pathway enrichment

At each selection p-value cutoff, we mapped positively and negatively selected contmeQTL SNPs to all genes overlapping a ±500 kb window around each SNP (hg19), excluding genes mapped to both sets, and tested each gene list for enrichment of GO, KEGG and Reactome terms using *gprofiler2*. For the most enriched terms that were reported in the main analysis, we calculated the proportion of selection cutoffs that each term was significantly enriched (adjusted p < 0.05), as well as the number of significant terms per cutoff.

Genetic risk for neropsychiatric and autoimmune traits

At each selection p-value cutoff, we tested whether positively selected contmeQTLs carried increased GWAS risk signal than positively selected non-contmeQTLs for a set of autoimmune and neuropsychiatric traits. For each variant, we computed the GWAS Z-score (β/SE), aligned to the responsive allele, for six genomic factors of neuropsychiatric disorders (a general psychopathology factor, compulsive disorders, schizophrenia and bipolar disorders, neurodevelopmental disorders, internalizing disorders, and substance use) and for type 1 diabetes and asthma. As a control trait, we included red hair colour. The neuropsychiatric GWASs had already undergone basic QC for use in GenomicSEM, and for the latter four traits, we filtered variants with MAF of at least 1%. We analyzed the reported test statistic for variants of interest to calculate the median Z-score across positively selected contmeQTLs. Null distributions were obtained by repeatedly drawing allele-frequency-stratified sets of the same size from the positively selected non-contmeQTLs within the allele frequency range of the observed set, and computing the median for the non-contmeQTLs. The empirical p-value was calculated as the proportion of permuted medians equal to or larger than the observed median.

### **Supplementary Results**

#### 1. Sex-specificity and distance between SNP and CpG

All contmeQTLs were sex-consistent, i.e., they showed the same effect direction in females and males. Furthermore, CpGs regulated by contmeQTLs neither overlapped with the 250 top marker CpG-sites nor with the 1,000 top marker CpG-sites for blood cell-types identified in Loyfer at al. ^53^, suggesting the absence of strong cell-type specific effects.

SNPs involved in the 5,090 *cis* contmeQTLs were significantly further apart from the respective CpG-site (mean absolute distance=201,680 bp) as compared to the distance between SNP and CpG-site of all tested non-significant combinations (mean absolute distance=97,807 bp, p < 2.2×10^−16^, Wilcoxon-test) indicating more long-range regulation within these contmeQTLs. There was no significant difference in the magnitude of effect size between SNPs located in close proximity to the respective CpG-site and more distal SNPs.

#### 2. Sensitivity analysis: including BMI and running the saturated model

We performed further sensitivity analyses on the significant contmeQTLs. As suggested by Keller ^56^, we ran a fully saturated model including the interaction term of every covariate with the respective SNP and CA. Furthermore, we included BMI as additional covariate. Interaction effects estimates were highly concordant with those from the primary analyses (ALSPAC: r=0.95, BioMD-Y: r=0.98). Furthermore, effect directions were highly consistent (ALSPAC: 96% of effect directions consistent, BioMD-Y: 99% of effect directions consistent).

#### 3. Enrichment for chromHMM states

To evaluate the potential gene regulatory function of contmeQTL SNPs and contmeQTL CpGs we performed enrichment analyses for chromHMM marks, based on ChIpSeq experiments in blood cell lines from the Roadmap project ^86^. We found significant enrichment for enhancers for contmeQTL CpGs (p=2.37×10^−12^, OR=1.25) as well as contmeQTL SNPs (p=1.05 × 10^−03^, OR=1.14) as compared to all tested SNPs and CpG-sites. ContmeQTL CpG-sites were additionally enriched for genic enhancers (p=3.06 × 10^−07^, OR=1.40) and flanking active transcription start sites (p=3.06×10^−14^, OR=1.27) and depleted for quiescent states (p=2.18×10^−07^, OR=0.86) and repressed weak PolyComb (p=1.67×10^−04^, OR=0.94, all Fisher-tests). This aligns with the previously reported relevance of enhancer regulation in complex trait genetics and environmental sensitivity ^87,88^.

#### 4. CpGs regulated by contmeQTLs overlap with GWAS and EWAS hits

The proportion of overlap with GWAS hits was significantly larger for contmeQTL SNPs (overlap of 44.7%, n=3,778 SNPs out of 8,425 unique contmeQTL SNPs) as compared to all tested meQTL SNPs (overlap of 36.8%, n=61,740 SNPs out of 167,600 unique tested SNPs; p=3.54x10^-23^, rate ratio=1.23, Poisson rate test). We not only found a higher degree of overlap, but also significant differences with regards to domains the GWAS hits mapped to: contmeQTL SNPs mapped significantly more often to the domains ‘Autoimmune immunological disease’ (e.g. “Rheumatoid arthritis”, rate ratio=2.08, p=5.42×10^−236^), ‘Endocrine’ (e.g. “Hypothyroidism”, rate ratio=2.15, p=1.15×10^−101^), ‘Gastrointestinal’ (e.g. “Diseases of the Digestive System”, rate ratio=1.36, p=3.58×10^-04^), ‘Haematological’ (e.g. “Red blood cell counts”, rate ratio=1.10, p=4.66×10^−28^), ‘HLA’ (e.g. “HLA DR on B cell”, rate ratio=2.70, p=9.48×10^−161^), ‘Mental Disorders’ (e.g. “Schizophrenia”, rate ratio=1.11, p=6.95×10^−03^), ‘Nutritional’ (e.g. “Fresh fruit intake”, rate ratio=1.83, p=3.80×10^−171^), ‘Pulmonary’ (e.g. “Lung function “FVC)”, rate ratio=1.28, p=2.43×10^−28^) and Surgical (e.g. “appendicectomy”, rate ratio=1.77, p=1.54×10^−33^, all Fisher’s enrichment tests) compared to all tested meQTL SNPs (see **Figure 1C**, **Tables S3** and **S4**). Furthermore, contmeQTL SNPs were not enriched for SNPs that had been associated with exposure to childhood maltreatment identified by Warrier et al. ^89^ nor for glucocorticoid receptor (GR)-response meQTLs ^64^.

While the relative proportion of CpG-sites overlapping with EWAS-hits from the EWAS catalogue did not significantly differ between all tested meQTLs CpGs and contmeQTL CpGs, they mapped to significantly different domains (see **Tables S5** and **S6**): contmeQTL CpGs mapped significantly more often to traits from the domains ‘Cardiovascular’ (e.g. “incident stroke”, rate ratio=1.96, p=7.74×10^−05^), ‘Gene expression’ (e.g. “PEX6”, rate ratio=2.51, p=3.09×10^−27^), ‘Sex’ (i.e. genetic sex, rate ratio=1.19, p=1.95×10^−09^), ‘Socioeconomic’ (e.g. “socioeconomic position”, rate ratio= 1.91, p= 5.52×10^−03^), ‘Smoking’ (e.g. “smoking current vs. never smoking”, rate ratio=1.36, p=3.10×10^−11^), ‘Maternal Smoking’ (e.g. “maternal smoking during pregnancy”, rate ratio=2.23, p=2.42×10^−18^), ‘Neurological’ (e.g. “Alzheimer’s disease”, rate ratio=1.62, p=1.56×10^−03^), all Poisson rate tests). Smoking and socioeconomic position are well-established correlates of blood DNA methylation ^90,91^. Because smoking is captured only imperfectly by covariate adjustment, including by methylation-based proxies, and smoking is associated with socioeconomic position, residual confounding cannot be excluded ^92,93^. We therefore specifically checked contmeQTL CpGs significantly enriched over meQTL CpGs for EWAS hits for “Maternal Smoking”, “Smoking” or “Socioeconomic”. Even though the enrichment was significant, only 6% (n=289 unique CpGs) of all contmeQTL CpGs overlapped with these categories and none of these CpGs were regulated by SNPs included in the PIRS (see Results).

#### 5. ContmeQTLs are enriched for pathways implicated in the major histocompatibility (MHC)-protein complex

Enrichment for GO terms highlighted several pathways implicated in the major histocompatibility (MHC)/human leukocyte antigen (HLA)-complex (located on chr6:28,477,797-33,448,354, based on hg19, see **Figure 1D**), besides other terms. Genes mapping to contmeQTL CpG-sites and contmeQTL SNPs were significantly enriched for the following GO terms: ‘MHC-protein-complex’ (CpGs: p=3.66×10^−09^, relative overlap=0.83, SNPs: p=2.03×10^−09^, relative overlap=0.88), ‘peptide antigen assembly with MHC-class-II-protein complex’ (CpGs: p=3.97×10^−08^, relative overlap=1.00, SNPs: p=8.00×10^−08^, relative overlap=1.00), ‘MHC-class-II-protein-complex’ (CpGs: p=3.65×10^−07^, relative overlap=0.91; SNPs: p=8.00×10^−08^, relative overlap=1.00), ‘peptide antigen assembly with MHC-protein complex’ (CpGs: p=3.91×10^−07^, relative overlap=0.85, no significant enrichment for SNPs), ‘luminal side of endoplasmic reticulum (ER) membrane’ (CpGs: p=2.14×10^−06^, relative overlap=0.68, SNPs: p=4.86×10^−06^, relative overlap=0.68), ‘peptide antigen binding’ (CpGs: p=6.70×10^−06^, relative overlap=0.61, SNPs: p=1.56×10^−05^, relative overlap=0.61), ‘lumenal side of membrane’ (CpGs: p=7.90×10^-05^, relative overlap=0.54, SNPs: p=1.56×10^−05^, relative overlap=0.68) and ‘MHC-class-II-protein-complex binding’ (CpGs: p=3.83×10^−05^, relative overlap=0.60, SNPs: p=1.72×10^−05^, relative overlap=0.67, all Fisher-tests). *HLA*-genes with the highest number of mapped contmeQTL CpGs were *HLA-J* (21 mapped CpG-sites) and *HLA-DBP2* (19 mapped CpGs) while *HLA-B* (100 mapped SNPs) and *HLA-J* (48 mapped SNPs) were the HLA-genes with the highest number of mapped contmeQTL SNPs. *HLA-J* has been mainly linked to cancer ^94-96^, and major associations for *HLA-B* have been reported for autoimmune disorders ^97,98^. *HLA-DBP2* and *HLA-B* have also been associated with mental disorders such as psychotic experiences ^99^, depression and anxiety ^100^. Other enriched terms in both contmeQTL SNPs and contmeQTL CpGs genes included terms related to cell-cell interaction and synapse (see **Figure 1D**).

#### 6. ContmeQTLs regulate gene expression

To evaluate if contmeQTL CpG-sites were also associated with changes in gene expression, we performed expression quantitative trait methylation (eQTM) analysis in the KORA-cohort. Here, DNAm as well as gene expression and information of exposure to CA were available for 1,519 individuals. From the overall 5,038 contmeQTLs CpGs available, about 30% (n=1,542 CpGs) were significantly associated with gene expression with up to twenty genes (average of 1.8 genes, see **Table S9**). To pick an illustrative example: the most significant contmeQTL, which was also an eQTM, was the CpG-site cg01376079 which was positively associated with the gene expression of *CLCF1* (beta=0.3539, p=1.63×10^−03^) and negatively associated with the gene expression of *SSH3* (beta=-1.2580, p=4.13×10^−34^). *CLCF1* is a cytokine belonging to the IL-6 family ^101^. *SSH3* has been linked to the notch signaling pathway ^102^ which plays a key role in inflammatory response ^103^. Cg01376079 is regulated by the contmeQTL SNP rs12789637 in interaction with CA (p=6.18×10^-08^). With regard to *HLA*-genes, 64 CpG-sites were significantly associated with gene-expression of *HLA-DRB5* (most significantly associated CpG cg11720950, beta=29.78, p=1.45×10^−56^) and 47 CpG-sites with the gene-expression of *HLA-DRB6* (most significantly associated CpG cg04418355, beta=-5.95, p=6.88×10^−99^). Both *HLA*-genes have been associated with a number of disorders, including narcolepsy, rheumatoid arthritis and multiple sclerosis ^104-106^ but also with schizophrenia and autism ^107^. From the 111 CpG-sites associated with *HLA-DRB5* or *HLA-DRB6*, 91 were also available in a sample of 179 individuals from the BeCOME cohort where DNAm as well as gene expression levels had been assessed. For the majority of these CpG-sites (82%), the direction of association between DNAm and gene expression levels where consistent with the respective eQTM in KORA. Furthermore, we could replicate the top eQTMs from KORA at FDR 0.05 (cg11720950 and *HLA-DRB5*: beta=20.36, p=1.08×10^−04^, cg04418355 and *HLA-DRB6:* beta=-3.79, p=2.14×10^−07^).

7. Overlap across different layers

Given that we assessed evidence for function and disease enrichment of contmeQTL CpGs and contmeQTL SNPs on different levels (overlap with EWAS and GWAS hits, enrichment for significant MR results and eQTMs), we investigated if any markers showed associations across these layers. Overall, 17 contmeQTL CpGs overlapped with eQTMs identified in KORA and had been potentially associated with complex traits in the MR-analysis by Richardson et al. ^63^. All of these also overlapped with EWAS hits and the respective contmeQTL SNPs overlapped with GWAS hits for complex traits (see **Figure S2** and **Table S10**). Looking more closely into these results reveals the complex mechanisms which might underlie contmeQTLs. For further illustration, we focus on rs3130724 (chr6: 29,118,530) located in the *MHC*-complex, interacting with CA on cg00765128 (chr6: 28,226,905), located in *ZKSCAN4*. Rs3130724 has been associated with a variety of GWAS-traits, including autoimmune disease and mental disorders, cg00765128 has been identified as potentially causally linked to autoimmune disorders in the MR-analysis. Furthermore, it is associated with the gene expression of *ZKSCAN4* in KORA (p=4.30×10^-05^, see **Figure S2**)*.* Reduced gene expression of *ZKSCAN4* has been found in the hippocampi of depressed patients and stress-susceptible mice ^108^ and the gene has been implicated in the shared genetic architecture of post-traumatic stress- and gastrointestinal disorders ^109^. Interestingly, rs3130724 has additionally been reported as blood expression quantitative trait (eQTL) associated with the expression of *HLA-K* in the GTEx project ^110^, adding another possible layer of regulation. Genetic variants in this pseudogene have been associated with immune response due to Covid-19 severity ^111^ and with autoimmune disorders ^112^.

#### 8. Pathway enrichment for contmeQTLs in childhood cohorts

For pathway enrichments, we used the CpGs from the 5,120 contmeQTLs identified in adolescent/adult cohorts as background to test if we observed enrichment on top of these. This revealed that the 891 unique genes mapping to CpGs regulated by the replicated 1,061contmeQTLs were even more enriched for ‘MHC-protein-complex’ (p=2.66×10^−06^, relative overlap=0.87), ‘MHC-classII-protein complex’ (p=6.31×10^−05^, relative overlap=0.90) as well as for ‘lumenal side of endoplasmic reticulum membrane’ (p=2.71×10^−05^, relative overlap=0.85) and ‘lumenal side of membrane (p=2.71×10^−05^, relative overlap=0.85)’. Furthermore, we observed significant enrichment for ‘antigen processing and presentation of exogenous peptide antigen’ (p=1.63×10^−06^, relative overlap=0.92) and ‘antigen processing and presentation of peptide antigen’ (p=4.18×10^−06^, relative overlap=0.74). All significant enrichments were driven by the overlap of *HLA*-genes with the specific pathways further strengthening the regulatory role of inflammation also in the childhood cohorts.

#### 9. Enrichment of contmeQTLs for positive selection

To support the enrichment of contmeQTLs for positive selection, we permuted 10,000 allele frequency and size-matched sets of LD-pruned non-contmeQTLs and computed, for each permutation, the proportion of variants meeting the same positive-selection threshold. This confirmed the enrichment: 0.6% of contmeQTLs met the positive-selection threshold, compared with a null mean of 0.226% (95% CI = 0.09 - 0.37%) among matched non-contmeQTLs (empirical p=0.0001).

#### 10. Survival advantage for PIRS+ variants

Since the PIRS+ was built from positively-selected variants, we tested whether a similar survival benefit would be expected for any comparable set of positively-selected regulatory variants. We repeated the survival analysis across 10,000 allele frequency and size-matched sets of positively-selected, LD-pruned non-contmeQTLs, substituting each matched set for the PIRS+ in the Cox model. As expected, positively-selected non-contmeQTLs were, on average, also associated with better survival (mean HR=0.92, 95% CI = 0.915 - 0.918, p < 2x10^-16^). However, the survival advantage was significantly more pronounced for contmeQTLs: the observed PIRS+ hazard ratio was more extreme than 97.3% of the 10,000 matched permutations (empirical p=0.027).

#### 11. Pathway and disease enrichment for positively-selected contmeQTLs

We tested which biological pathways were linked to selection at contmeQTLs. Genes near positively-selected contmeQTLs were broadly enriched for immune-related pathways: 17 / 19 significantly enriched terms were immune or autoimmune disease-related (see **Figure S3A**). Notable enrichments included Th1 and Th2 cell differentiation (adjusted p=2×10⁻^0^⁴), Toll-like receptor 2 binding (adjusted p=0.003), and the MHC class II protein complex (adjusted p=0.013). None of these terms were enriched among genes near negatively-selected contmeQTLs or near selected non-contextual meQTLs, indicating the enrichment is more specific to positively-selected, contextual meQTLs rather than a general feature of meQTLs or of selection itself.

Regarding disease enrichment, the strongest observed enrichments were for autoimmune disease terms, including asthma (adjusted p=4×10⁻^06^). We tested this relative to positively-selected non-contmeQTLs across 10,000 allele frequency and size-matched permutations, the median asthma GWAS risk was significantly higher in PIRS+ alleles (Z = 0.46, permutation p = 0.0095), as was type 1 diabetes risk (Z = 0.36, p = 0.016). Among neuropsychiatric traits, PIRS+ alleles showed elevated risk for the psychosis (F2; Z = 0.34, p < 0.0001) and substance use (F5; Z = 0.30, p < 0.0001) genomic factors by ^84^, but not for the internalizing (F4), compulsive (F1), or general psychopathology (P-factor) factors, and there was no enrichment or depletion for the negative-control trait red hair (see **Figure S3B**). Together, these results indicate that recent positive selection has acted on contmeQTLs that elevate genetic risk for autoimmune disease and specific psychiatric conditions.

#### 12. Sensitivity evolutionary analyses

###### Assessing the robustness of the observed enrichment for positive selection in contmeQTLs

Across the 14 selection p-value cutoffs tested, ranging from 1×10^‑08^ to 0.05, the number of positively selected contmeQTLs increased from 14 to 340 LD-pruned variants (**Figure S4A**). Originally, we had used the FDR cutoff for genome-wide significance as presented in Akbari et al.^75^, resulting in 18 variants. ContmeQTLs were enriched for positive selection relative to non-contmeQTLs at every selection p-value cutoff ≤ 1×10^‑04^ with the Firth OR ranging from 3.78 (95% CI: 1.89-7.17, p = 3.2×10^‑04^) at 1×10^‑08^ to 1.58 (95% CI: 1.19-2.08, p = 0.002) at 1×10^‑04^ (**Figure S4B**). The permutation test confirmed these enrichments, with significant enrichment for positively selected contmeQTLs relative to non-contmeQTLs at every cutoff ≤ 1×10^‑04^ (empirical p ≤ 1×10^‑04^). At 5×10^‑04^, the permutation test remained nominally significant (empirical p = 0.036), but the Firth 95% CI included 1. For all subsequent analyses, we therefore focused on p-value cutoffs that showed significant enrichment of contmeQTLs for positively selected alleles across both the Firth regression and permutation methods in order to ensure that reported associations reflect variants with evidence of positive selection, rather than to analyze sets of contmeQTLs that likely include an increasing proportion of weakly selected or selection false-positives.

###### Robustness of the PIRS+ association with survival in ancient individuals

The PIRS+ computed across selection p-value thresholds used 14 to 62 SNPs across the cutoffs. For the samples in the top-quartile of PIRS+, the HR estimate was below 1 at every cutoff up to 1×10^‑05^ (HR = 0.80-0.94; **Figure S4C**), but was only significantly less than 1 at the three most stringent cutoffs (1×10^‑08^: HR = 0.80, 95% CI: 0.67-0.96; 5×10^‑08^ and 1×10^‑07^, identical 15-SNP score: HR = 0.82, 95% CI: 0.68-0.99). At these cutoffs, PIRS+ also outperformed scores built from matched, positively selected non-contmeQTLs (empirical p = 0.0089, 0.027 and 0.043). The association attenuated as more weakly selected variants were included (HR = 0.85-0.94 between 5×10^‑07^ and 1×10^‑05^; empirical p ≥ 0.17) and the HR estimate was greater than 1 (but not statistically significant) at 5×10^‑05^ and 1×10^‑04^ (HR = 1.08 and 1.12). Overall, the survival advantage of PIRS+ appears to be driven by the most strongly selected contmeQTLs with no association at weaker selection thresholds.

###### Robustness of the gene pathway enrichments.

Across the sensitivity tests of gene pathways enriched near positively selected contmeQTLs, the broad enrichment for immune-related genes was stable across selection p-value cutoffs. Notably, the KEGG asthma pathway ranked first or second at every cutoff from 5×10^‑08^ to 1×10^‑04^. Of the most strongly enriched terms in the main analysis, 11 were significantly enriched at 5 or more of the 9 cutoffs ≤ 1×10^‑04^: keratohyalin granule at all nine; asthma, inflammatory bowel disease, Toll-like receptor 2 binding, intestinal immune network for IgA production, Th1 and Th2 cell differentiation and allograft rejection at eight; autoimmune thyroid disease at seven; and Th17 cell differentiation and leishmaniasis at six.

###### Robustness of GWAS risk enrichment at positively selected contmeQTLs

GWAS risk enrichment for the neuropsychiatric factors could only be tested at six selection p-value cutoffs, ranging from 5×10^‑07^ to 1×10^‑04^. At more stringent selection cutoffs, the matched null pool was too small to run the permutation testing (at these cutoffs, fewer positively selected non-contmeQTLs with GWAS summary statistics fell within the allele frequency range of the observed set than there were positively selected contmeQTLs, making permutation testing unreasonable). Across the neuropsychiatric GWAS traits examined with adequate data, the elevated substance use risk was the most stable enrichment signal, significant at all six cutoffs (median responsive-allele Z across positively selected contmeQTLs always > 0, and all empirical p-values < 0.02) (Figure S5). Elevated schizophrenia-bipolar disorder (psychosis) risk was only observed at cutoffs of 5×10^‑06^ or below (median Z = 0.31-0.34, p ≤ 0.018), and the median Z for neurodevelopmental disorders was negative at all six cutoffs (‑0.23 to ‑0.46). Among the autoimmune traits, type 1 diabetes risk was elevated at all cutoffs (median Z = 0.27-0.44, all p-values < 0.025). Asthma risk was highest among the most strongly selected variants (median Z = 1.14, p < 1×10^‑04^ at selection p-value < 1×10^‑08^) and attenuated thereafter.

### Supplementary Figures

#### Figure S1: Overview of statistical analyses

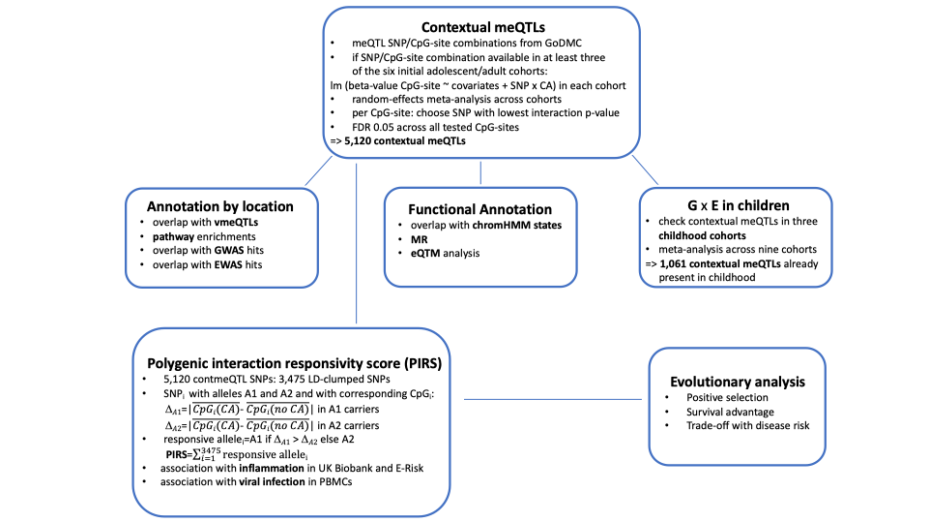

#### Figure S2: Overlap across different layers

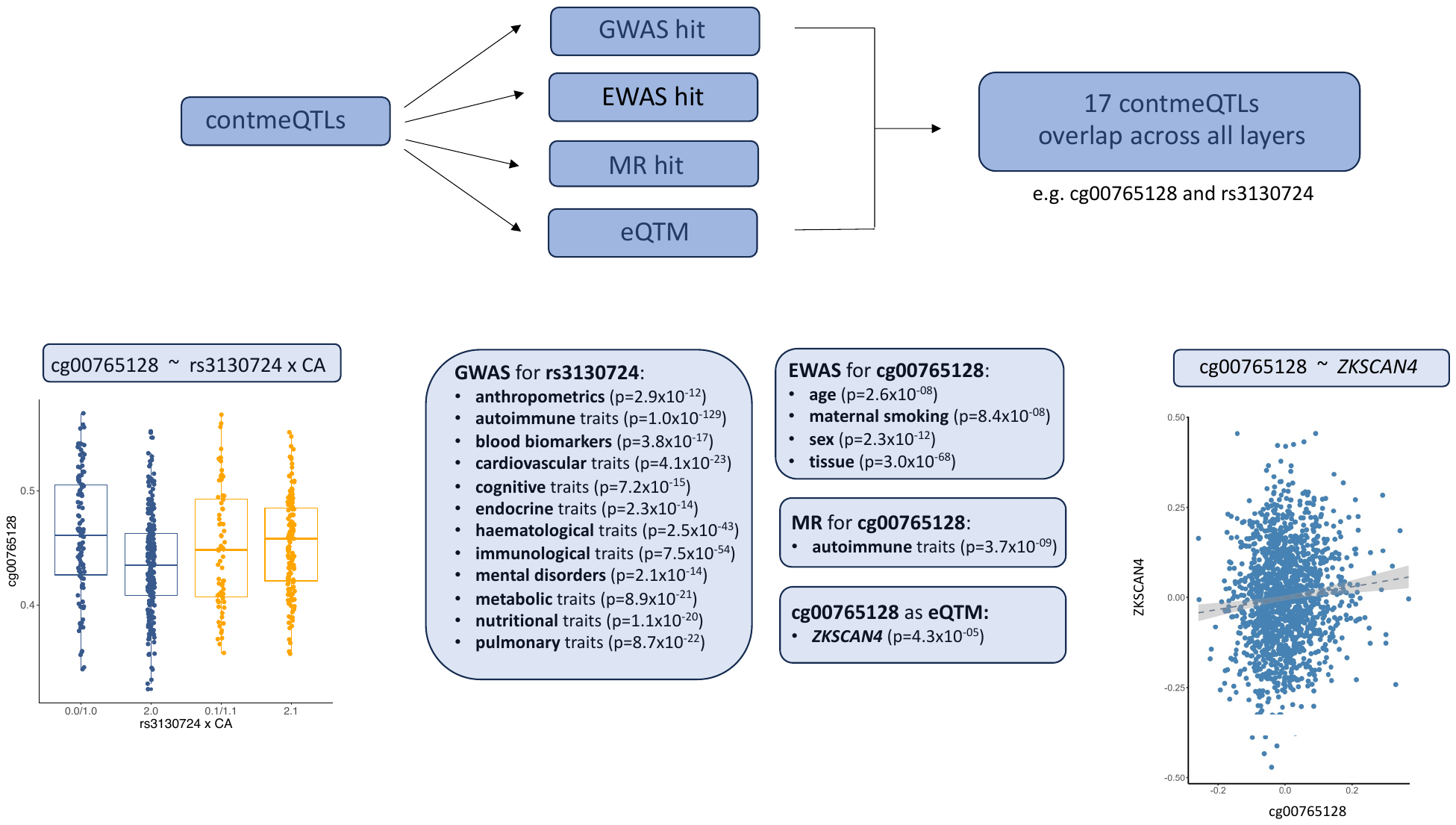

#### Figure S3: PIRS+ pathway enrichments

(**A**) Enrichment for PIRS+ variants, as compared to non-contmeQTLs, for KEGG pathways, GO terms and Reactome pathways

(**B**) Enrichment for PIRS+ variants for GWAS traits

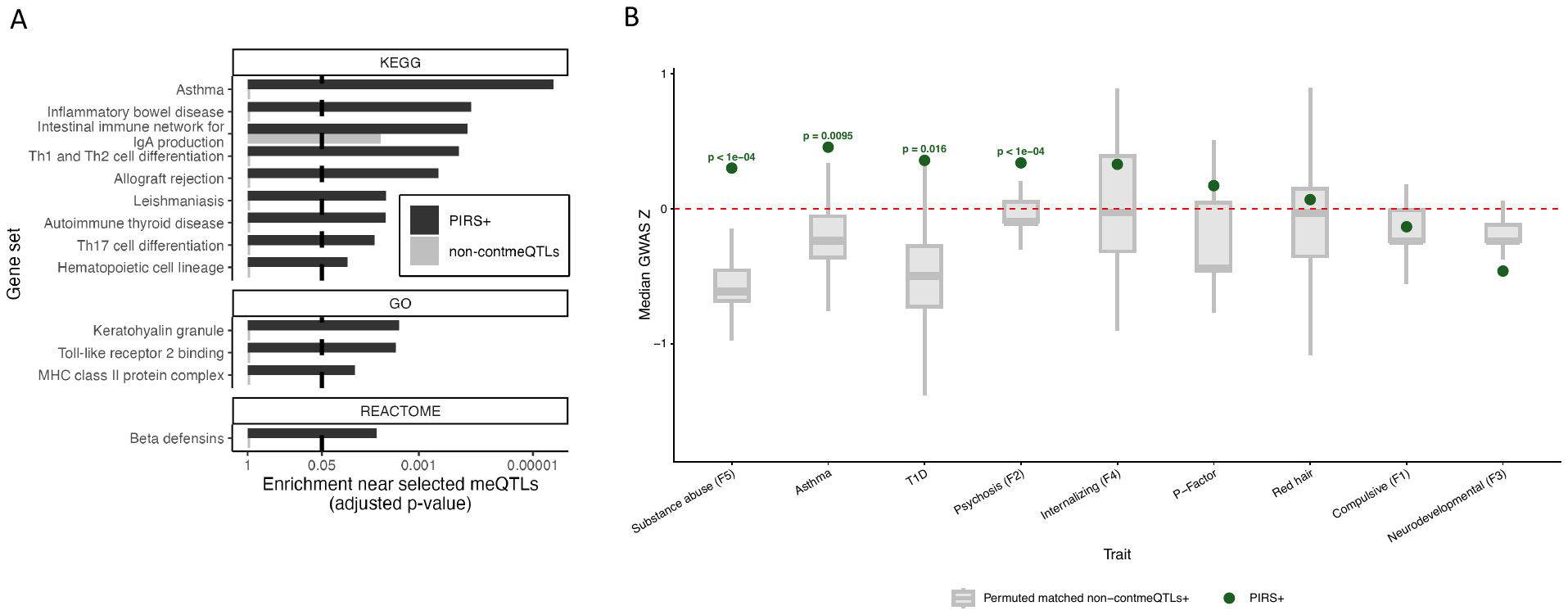

#### Figure S4: Sensitivity analyses on positive selection thresholds

(**A**) Number of positively selected contmeQTLs dependent on selection p-value cutoff, the red bar indicates the number of variants included in the initial PIRS+

(**B**) Enrichment of contmeQTLs for positive selection dependent on selection p-value cutoff

(**C**) Survival HR for PIRs+ variants dependent on selection p-value cutoff

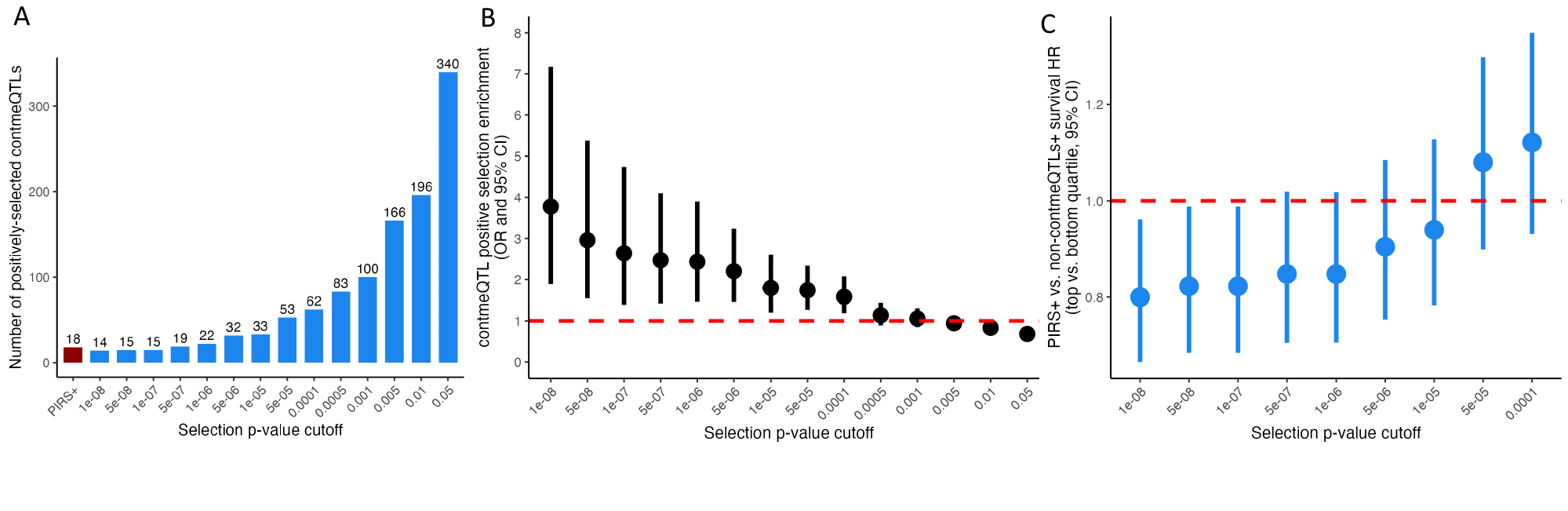

### Cohort specific study groups

#### 1. BeCOME study group

The members of the BeCOME study group are:

Elisabeth B. Binder^1^, Tanja Brückl^1^, Victor I. Spoormaker^1^, Angelika Erhardt^1^, Susanne Lucae^2^, Philipp G. Saemann^2^, Norma C. Grandi^1^, Tamara Namendorf^2^, Michael Czisch^2^, Immanuel Elbau^3^, Laura Leuchs^1^, Anna K. Brehm^4^, Leonhard Schilbach^5^, Julia Fietz^1^, Sanja Ilić-Ćoćić^1^, Julius Ziebula^2^, Iven-Alex von Mücke-Heim^1^, Yeho Kim^2^, Julius Pape^2^, Kerstin Hupe^2^, Michael Gottschalk^2^, Alexandros Balaskas^2^

1 Max Planck Institute of Psychiatry, Department Genes and Environment, Munich, Germany
2 Max Planck Institute of Psychiatry, Munich, Germany
3 Psychiatry Department, Weill Cornell Medicine, NY, USA
4 University Hospital of Old Age Psychiatry, University of Bern, Bern, Switzerland
5 LVR-Klinikum Düsseldorf - Kliniken der Heinrich-Heine-Universität, Düsseldorf, Germany

#### 2. Optima study group

The members of the OPTIMA study group are:

Johannes Kopf-Beck^1,2^, Samy Egli^3^, Martin Rein^4^, Nils Rek^4^, Julia Fietz^1^, Jeanette Tamm^2^, Katharina Rek^4^, Martin E. Keck^5^

1 Max Planck Institute of Psychiatry, Department Genes and Environment, Munich, Germany
2 Department of Psychology, Ludwig Maximilian University of Munich, Munich, Germany
3 Max Planck Institute of Psychiatry, Department Clinical Translation, Munich, Germany
4 Max Planck Institute of Psychiatry, Munich, Germany
5 Schmieder Hospital in Gailingen, Gailingen, Germany
